# Genetic Architecture And Clinical Outcomes Of The Fredrickson-Levy-Lees Dyslipoproteinemias

**DOI:** 10.1101/2022.05.26.22275391

**Authors:** Thomas Gilliland, Jaqueline S. Dron, Margaret Sunitha Selvaraj, Mark Trinder, Kaavya Paruchuri, Sarah M. Urbut, Sara Haidermota, Rachel Bernardo, Md Mesbah Uddin, Michael C. Honigberg, Gina Peloso, Pradeep Natarajan

## Abstract

**Background and Aims:** The genetic basis and clinical relevance of the classical Fredrickson-Levy-Lees (FLL) dyslipoproteinemia classifications has not been studied in general population-based cohorts. We aimed to evaluate the phenotypic and genetic characteristics of FLL disorders.

**Methods:** Among UK Biobank participants free of prevalent coronary artery disease (CAD), we used blood lipids and apolipoprotein B concentrations to infer FLL classes (Types I, IIa, IIb, III, IV, and V). For each FLL class, Cox proportional hazards regression estimated risk of incident CAD. Phenome-wide association testing was performed. GWAS were performed, followed by *in silico* causal gene prioritization and heritability analyses. Prevalence of disruptive Mendelian lipid variants was assessed from whole exome sequencing.

**Results:** Of 450,636 individuals, 259,289 (57.5%) met criteria for a FLL dyslipoproteinemia: 63 (0.01%) type I; 40,005 (8.9%) type IIa; 94,785 (21.0%) type IIb; 13,998 (3.1%) type III; 110,389 (24.5%) type IV; and 49 (0.01%) type V. Over median 11.1 years follow-up, compared to normolipidemics the type IIb pattern conferred the highest hazard of incident CAD overall (HR 1.92, 95% CI 1.84-2.01, *P*<0.001) and in meta-analysis across matched non-HDL-C strata (HR 1.45, 95% CI 1.30-1.60). GWAS revealed 250 loci associated with FLL, of which 13 were shared across all classes; compared to GWAS of isolated lipid traits, 72 additional loci were detected. Mendelian lipid variants were rare (2%), but polygenic heritability was high, ranging from 23% (type III) to 54% (type IIb).

**Conclusions:** FLL classes have distinct genetic architectures yielding new insights for cardiometabolic disease beyond single lipid analyses.

## Introduction

Over 50 years ago, Fredrickson, Levy, and Lees (FLL) introduced a classification scheme for lipid disorders based on patterns of circulating lipoproteins(1). In their seminal series of papers, the authors describe six dyslipoproteinemia types involving excess of one or more lipoprotein fractions. These were termed types I, IIa, IIb, III, IV, and V, reflecting chylomicron (CM) excess, low-density lipoprotein (LDL) excess, combined LDL and very low-density lipoprotein (VLDL) excess, remnant cholesterol-rich lipoprotein excess, VLDL excess, and combined CM and VLDL excess, respectively (**Table 1**). The authors observed distinguishing clinical characteristics and patterns of heritability for each disorder and hypothesized pathophysiologic mechanisms.

**Table 1.** Definition and characteristics of FLL dyslipoproteinemias. FLL disorders were defined as previously based on apoB and TG and the ratios of TG/apoB and TC/apoB. ApoB and TG are expressed in mg/dL for both isolated measurements and in defining the thresholds for the ratios TG/apoB and TC/apoB. Lipoprotein excess refers to the lipoprotein particle(s) found in excess for each phenotype. Abbreviations: ‘apoB’ = apolipoprotein B; ‘TG’ = triglycerides; ‘TC’ = total cholesterol; ‘LDL’ = low-density lipoprotein; ‘VLDL’ = very low-density lipoprotein.

| Fredrickson-Levy-Lees class definitions |  |  |  |  |  |
| --- | --- | --- | --- | --- | --- |
|  | ApoB (mg/dL) | TG (mg/dL) | TG/apoB | TC (mg/dL) /apoB | Lipoprotein excess |
| **Type I** | <75 | $\geq 133$ | $\geq 8.8$ | - | Chylomicrons |
| **Type IIa** | $\geq 120$ | < 133 | - | - | LDL |
| **Type IIb** | $\geq 120$ | $\geq 133$ | - | - | LDL and VLDL |
| **Type III** | < 120 | $\geq 133$ | < 8.8 | $\geq 2.4$ | Remnant particles |
| **Type IV** | < 120 | $\geq 133$ | < 8.8 | < 2.4 | VLDL |
| **Type V** | $\geq 75$ and < 120 | $\geq 133$ | $\geq 8.8$ | - | Chylomicrons and VLDL |

Owing to the cost and complexity of laboratory techniques used for diagnosis, the FLL phenotypes have been studied primarily in specialized lipid clinics and research cohorts. These settings may tend to capture extreme phenotypes, and conclusions drawn may not generalize to contemporary general populations beset with growing obesity and diabetes epidemics. Thus, the proposed clinical rationale for FLL classification beyond considering conventional lipid measures alone remains theoretical. Sniderman and colleagues developed and validated a diagnostic algorithm for the FLL disorders based on serum total cholesterol (TC), triglycerides (TG), and apolipoprotein B (apoB)(2), allowing FLL diagnosis with routine lipid measurements. Application to contemporary Western populations reveals a high prevalence of FLL dyslipoproteinemia with relationships to subclinical atherosclerosis(3,4). Associations with clinical outcomes in large contemporary cohorts are presently unknown.

Monogenic factors have largely been studied as the basis of FLL classes but comprise a very small fraction of individuals with dyslipoproteinemias. Inheritance patterns for FLL disorders are believed to be variably monogenic, oligogenic, and polygenic, but characterizations have largely been restricted to the former. Furthermore, phenotypic expression may depend on the interactions between genetic and environmental factors(5),(6). Expression of the type III pattern in patients with *APOE2* homozygosity is modified by mutations in other lipid-related genes(7,8) and likely requires comorbid disease or exposure – hyperinsulinemia, alcohol excess, or the postmenopausal state(9). Further, clinical type III can occur independently of *APOE2*(10). While the type II phenotype with isolated hypercholesterolemia (type IIa) is classically monogenic (i.e., familial hypercholesterolemia [FH]), FH represents a very small fraction of type II pattern(11,12). FLL type IIb is believed to be polygenic, but the genetic factors are not well understood(6),(13).

Here we apply the apoB-based classification scheme to a general population-based cohort to evaluate the clinical characteristics and cardiovascular outcomes of the FLL phenotypes. We complement our findings with genome-wide association studies of the FLL disorders and perform unbiased phenotype association testing to glean novel insights into lipid metabolic disturbances.

## Methods

### Study participants

The UK Biobank (UKBB) is a cohort of ~500,000 volunteer participants living in the United Kingdom at ages 40-69 years recruited across 22 sites between 2006 and 2010, with ongoing follow up(14). Extensive genetic and phenotypic data is available as previously described. Laboratory measurements, anthropometrics, and clinical histories were ascertained at study enrollment. This research was conducted under UK Biobank application number 7089. All study participants provided written and informed consent. The Mass General Brigham Institutional Review Board approved secondary use of these data (2013P001840).

### Biochemical measurement and definition of Fredrickson-Levy-Lees classes

Among UKBB participants, non-fasting blood lipids and apoB were measured at baseline. ApoB was quantified by immuno-turbimetric assay, TC and TG by enzymatic assay, high-density lipoprotein cholesterol (HDL-C) by enzyme immune-inhibition assay, and direct low-density lipoprotein cholesterol (LDL-C) by enzymatic selective protection assay, all using the Beckman Coulter AU5800 analytical platform, as previously reported(15). The presence of lipid-lowering medications was ascertained at baseline. Similar to prior studies, in the presence of a statin prescription, total cholesterol (TC) was divided by 0.8 and apoB and LDL-C were divided by 0.7(16,17).

Among 450,636 UKBB participants free of CAD at baseline, FLL phenotype was determined by an algorithm developed by Sniderman and colleagues, which utilizes apoB, TG and the ratios of TG/apoB and TC/apoB to group individuals into one of six dyslipoproteinemias (**Table 1**)(2). Individuals not meeting criteria for FLL were categorized as normolipidemic. Sensitivity analyses were performed among individuals who were not prescribed lipid-lowering medications (including statins, ezetimibe, fibrates, or niacin) (N = 382,159).

### Genotyping

UKBB samples were genotyped using the UK Biobank Lung Exome Variant Evaluation (UK BiLEVE) or Applied Biosystems UK Biobank Axiom Array. Genetic data was imputed using either the Haplotype Reference Consortium panel or the UK10K + 1000 Genomes panel(14,18).

### Clinical phenotypes

UKBB study subjects were free of CAD at the time of enrollment. The CAD definition was based on the appearance of a qualifying International Classification of Diseases (ICD) code corresponding to acute myocardial infarction and coronary artery revascularization, physician, or patient report, as used previously (**Supplementary Table 1**)(19–21). Similarly, the covariates of hypertension and type 2 diabetes were based on a combination qualifying ICD code or physician or patient report.

We coded 1584 phenome-wide phenotypes in UKBB from both prevalent and incident hospital episodes using ICD-9 and −10 diagnosis codes grouped into phecodes(22–24). Phecodes that were sex-specific were coded as not applicable for the opposite sex. Clinical phenotypes were defined using the Phecode Map 1.2 ICD-9 and ICD-10 phenotype groupings(25). ICD codes were queried up to March 2020 in UKBB.

### Monogenic dyslipidemia variant annotation

Monogenic dyslipidemia variant identification was performed on the subset of ~200,000 whole exome sequences (WES) available for UKBB participants who were also included in our study(26).

We extracted variants disrupting genes associated with monogenic dyslipidemia phenotypes, including high LDL-C, low HDL-C, and high TG. Each gene was defined on whether the resultant phenotype followed an autosomal dominant (AD) or autosomal recessive (AR) inheritance pattern. High LDL-C genes included *LDLR* (AD), *APOB* (AD), *PCSK9* (AD), *LDLRAP1* (AR), *ABCG5* (AR), and *ABCG8* (AR). Low HDL-C genes included: *APOA1* (AD), *ABCA1* (AR), and *LCAT* (AR). High TG genes included: *LPL* (AD), *APOA5* (AD), *APOC2* (AR), *GPIBHP1* (AR), *LMF1* (AR), and *APOE* (AR). Individuals were considered as having a consistent Mendelian lipid variant if they were either homozygous or heterozygous for the alternative allele in AD genes, or homozygous for the alternative allele in AR genes.

Variants with minor allele frequencies of <1% and <10% were considered in AD and AR genes, respectively. This list was further restricted based on multiple variant annotations; variants were included if they were defined as high-confidence loss-of function by the Ensemble Variant Effect Predictor (VEP) LOFTEE plugin, had a ClinVar interpretation of “pathogenic” or “pathogenic/likely pathogenic” (last checked November 2021), or were predicted as “damaging” by metaSVM(27–29). All variants were annotated following canonical gene transcripts.

Further, we determined genotypes for *APOE* (E2/E3/E4) based on allelic combinations from rs429358 and rs7412 (where rs429358 and rs7412 are T and T, respectively, for *APOE* E2; T and C for *APOE* E3; and C and C for *APOE* E4).

We used PLINK (version 2.0)(30) to perform variant extractions.

### Statistical analysis

For descriptive analyses, categorical variables are presented as frequencies and proportions and compared using chi square or Fisher’s exact test, and continuous variables are presented as mean with standard deviation or median with interquartile range and compared with Student’s T-test or Wilcoxon rank sum test.

In UKBB, incident CAD was compared between FLL phenotypes using Cox proportional hazards regression (‘survival’ R software package, version 3.2-13) relative to normolipidemic individuals. The model accounted for study enrollment to March 31^st^, 2020. Participants were censored if loss-to-follow-up or death occurred prior to March 31^st^, 2020. Covariates included age, sex, smoking status, genotyping array, the first five principal components of genetic ancestry, hypertension, type 2 diabetes, BMI, and statin prescription. In secondary analysis, individuals were sub-grouped according to non-HDL-C concentration (130-160 mg/dl, 160-190 mg/dl, 190-220 mg/dl, and 220-250 mg/dl); individuals with FLL types IIa, IIb, III, and IV were compared separately against a reference group of stratum-matched normolipidemic individuals for incident CAD using Cox proportional hazards regression. Heterogeneity testing was used to assess significant differences within non-HDL-C groups, and meta-analysis was performed across non-HDL-C groups to calculate an overall FLL phenotype effect (‘meta’ R software package, version 4.19-0)(31). Statistical significance was assigned at *P*<0.05.

We tested the phenome-wide associations between FLL disorders and combined prevalent and incident hospital episode statistic phenotypes using the PheWAS version 0.99.5-4 package for R (version 4.0.2). In a single model with normolipidemics as reference, the association between FLL class and each phecode was assessed using logistic regression with adjustments for age, sex, genotyping array, and the first 5 principal components of genetic ancestry. Accounting for multiple hypothesis-testing, statistical significance was assigned at *P*<3.2×10^−5^ (0.05/1584 phecodes).

We performed genome-wide association studies (GWAS) using REGENIE software (version 1.0.2) across four FLL phenotypes against normolipidemic controls(32). Third-degree related individuals (N=34828) were excluded by the KING method(33). REGENIE step 1 was performed on UKBB array genotypes. We excluded variants with minor allele frequency (MAF) <1%, minor allele count <100, genotype missingness >10%, or Hardy-Weinberg equilibrium p-value >10^−15^. Additionally, 6 samples (of 488377 used in step 1) were excluded due to genotype missingness >10%. In REGENIE step 2, Logistic regression was performed on genotyped and imputed variants (N=19475096) meeting MAF and imputation quality filters (MAF >=0.01% and INFO >=0.3), with Firth correction applied to SNPs where association p-value from standard logistic regression was <0.01. Covariates included were age, sex, genotyping array, and the first 10 principal components of ancestry. SNPs and indels with association p-values <5×10^−8^ were considered statistically significant. In a secondary analysis examining the effect of fasting status, we repeated the above GWAS additionally adjusting for fasting time, restricted to the top independent SNPs for each phenotype. Using LD score regression version 1.01 (restricted to SNPs present in HapMap3), we assessed the SNP heritability and pair-wise genetic correlation of FLL types IIa, IIb, III, and IV(34,35). We utilized a newly developed similarity-based method for gene prioritization, Polygenic Priority Score (PoPS), to investigate protein-coding gene contributions to each FLL phenotype(36). PoPS was performed on summary level GWAS data for each phenotype using 1000 Genome Project as a reference(37) to yield a ranked set of candidate causal genes.

Analyses were performed using R version 4.0.2 (R Core Team, 2020) unless otherwise specified.

## Results

### Baseline characteristics of UKBB Participants

Overall, 450,636 UKBB participants with non-missing apoB and lipids, free of prevalent CAD, and with active participant consent were included in the analysis. Mean age was 56.3 (8.1) years, 44.4% were male, and 94.3% were white (**Table 2)**. FLL dyslipoproteinemia was common, with 259,289 (57.5%) participants meeting criteria for one of the FLL types. The most prevalent FLL type was type IV (110,389, 24.5% of the overall population), followed by type IIb (94,785, 21.0%), type IIa (40,005, 8.9%), type I (63, 0.01%), and type V (49, 0.01%) (**Supplementary Figure 1**). Mean BMI was lowest among participants without dyslipoproteinemia (25.9 kg/m^2^), intermediate in type IIa individuals (26.9 kg/m^2^), and highest in FLL types I, IIb, IV, and V (28.9, 28.9, 28.8, and 29.3 kg/m^2^, respectively). Type 2 diabetes was more common in among individuals with FLL disorders I, IIb, IV, and V – 9.5%, 2.5%, 3.3%, and 10.2%, respectively – compared to normolipidemics or FLL disorders IIa and III – 1.4%, 1.2%, and 1.2%, respectively. Statin treatment was most common in participants with FLL types IIa and IIb (25.1% and 24.3%, respectively), compared to 8.3% of normolipidemics. The distribution of lipids adjusted for lipid-lowering treatment (see Methods) reflected the assignments according to the apoB-based algorithm (**Figure 1**).

**Table 2.** Baseline characteristics of UKBB study participants. We report demographic characteristics, baseline lipids, lipid-lowering medication use, and relevant medical history for all study participants (‘All’), individuals not meeting criteria for an FLL disorder (‘Normolipidemic’), and individuals meeting criteria for any of the six FLL dyslipoproteinemias. Abbreviations: ‘SD’ = standard deviation; ‘IQR’ = interquartile range; ‘TC’ = total cholesterol; ‘LDL-C’ = low density lipoprotein cholesterol; ‘HDL-C’ = high density lipoprotein cholesterol; ‘TG’ = triglyceride; ‘ApoB’ = apolipoprotein B; ‘BMI’ = body-mass index.

|  | <u>All</u> | <u>Normolipidemic</u> | <u>Type I</u> | <u>Type IIa</u> | <u>Type IIb</u> | <u>Type III</u> | <u>Type IV</u> | <u>Type V</u> |
| --- | --- | --- | --- | --- | --- | --- | --- | --- |
| **Number (%)** | 450636<br>(100) | 191347 (42.5) | 63 (0.01) | 40005<br>(8.9) | 94785<br>(21.0) | 13998<br>(3.1) | 110389<br>(24.5) | 49 (0.01) |
| **Demographics** |  |  |  |  |  |  |  |  |
| **Age at enrollment, mean (SD), y** | 56.3 (8.1) | 55.0 (8.3) | 55.8 (8.6) | 58.4 (7.3) | 57.9 (7.5) | 56.8 (7.7) | 56.6 (8.1) | 52.6 (8.6) |
| **Male sex, n (%)** | 200353<br>(44.4) | 67992 (35.5) | 57 (90.5) | 14991<br>(37.5) | 48317<br>(51.0) | 4792<br>(34.2) | 64163<br>(58.1) | 41 (83.7) |
| **White, n (%)** | 424851<br>(94.3) | 179080 (93.6) | 59 (93.7) | 37725<br>(94.3) | 90518<br>(95.5) | 13408<br>(95.8) | 104015<br>(94.2) | 46 (93.9) |
| **Black, n (%)** | 7114 (1.6) | 4635 (2.4) | 0 | 847 (2.1) | 635 (0.7) | 128 (0.9) | 869 (0.8) | 0 |
| **Baseline lipids** |  |  |  |  |  |  |  |  |
| **TC, median (IQR), mg/dl** | 225.8<br>(199.5-253.9) | 208.4 (186.7-229.6) | 176.4<br>(155.8-200.2) | 265.5<br>(247.1-285.7) | 267.4<br>(248.2-289.9) | 221.6<br>(197.1-247.0) | 214.5<br>(195.0-232.5) | 230.4<br>(206.8-256.5) |
| **LDL-C, median(IQR), mg/dl** | 143.0<br>(122.7-165.2) | 127.6 (111.6-142.9) | 90.5<br>(76.4-98.6) | 176.2<br>(166.0-189.5) | 178.5<br>(166.2-194.7) | 126.1<br>(109.8-143.1) | 137.2<br>(123.5-149.2) | 114.2<br>(107.0-129.6) |
| **HDL-C, median (IQR), mg/dl** | 54.5<br>(45.7-65.1) | 60.8 (51.5-71.2) | 32.7<br>(27.7-39.0) | 60.6<br>(51.9-70.7) | 49.5<br>(43.0-57.2) | 64.4<br>(54.6-74.1) | 46.9<br>(40.3-54.5) | 32.6<br>(29.5-37.4) |
| **TG, median (IQR), mg/dl** | 130.6<br>(92.3-188.9) | 90.3 (72.1-109.4) | 605.7<br>(512.2-696.1) | 105.1<br>(88.6-119.5) | 205.0<br>(165.5-267.6) | 179.3<br>(151.2-235.5) | 181.9<br>(153.6-232.7) | 839.3<br>(786.0-903.3) |
| **ApoB, median (IQR), mg/dl** | 107.3<br>(91.9-123.7) | 95.0 (83.5-106.0) | 57.4<br>(48.4-68.6) | 129.9<br>(124.2-139.7) | 135.4<br>(126.7-148.3) | 86.2<br>(75.0-97.1) | 104.7<br>(94.8-112.5) | 84.2<br>(78.4-94.3) |
| **Lipid-lowering therapy** |  |  |  |  |  |  |  |  |
| **Statin, n (%)** | 62990<br>(14.0) | 15903 (8.3) | 5 (7.9) | 9984<br>(25.0) | 22888<br>(24.1) | 522 (3.7) | 13680<br>(12.4) | 8 (16.3) |
| **Ezetimibe, n (%)** | 1861 (0.4) | 437 (0.2) | 1 (1.6) | 175 (0.4) | 623 (0.6) | 33 (0.2) | 592 (0.5) | 0 |
| **Fibrate, n (%)** | 1024 (0.2) | 245 (0.1) | 2 (3.2) | 57 (0.1) | 333 (0.4) | 30 (0.2) | 357 (0.3) | 0 |
| **Niacin, n (%)** | 39 | 14 | 0 (0%) | 1 | 11 | 2 | 11 | 0 (0%) |
| **Any therapy, n (%)** | 78477<br>(17.4) | 22718 (11.8) | 9 (14.3) | 11393<br>(28.5) | 25869<br>(27.3) | 1054 (7.5) | 17426<br>(15.8) | 8 (16.3) |
| **Medical history** |  |  |  |  |  |  |  |  |
| **Smoking status, n (%)** | 199707<br>(44.3) | 77564 (40.5) | 36 (57.1) | 17689<br>(44.2) | 46826<br>(49.4) | 6353<br>(45.4) | 51207<br>(46.4) | 32 (65.3) |
| Never | 248706<br>(55.2) | 112920 (59.0) | 27 (42.9) | 22145<br>(55.4) | 47466<br>(50.1) | 7573<br>(54.1) | 58559<br>(53.0) | 16 (32.7) |
| Previous | 152861<br>(33.9) | 60920 (31.8) | 27 (42.9) | 13759<br>(34.4) | 34631<br>(36.5) | 4920<br>(35.1) | 38581<br>(35.0) | 23 (46.9) |
| Current | 46846<br>(10.4) | 16644 (8.7) | 9 (14.3) | 3930 (9.8) | 12195<br>(12.9) | 1433<br>(10.2) | 12626<br>(11.4) | 9 (18.4) |
| **Hypertension, n (%)** | 126092<br>(28.0) | 41155 (21.5) | 30 (47.6) | 11699<br>(29.2) | 33336<br>(35.2) | 3573<br>(25.5) | 36279<br>(32.9) | 20 (40.8) |
| **Diabetes mellitus, type 2, n (%)** | 9316 (2.1) | 2624 (1.4) | 6 (9.5) | 467 (1.2) | 2366 (2.5) | 174 (1.2) | 3674 (3.3) | 5 (10.2) |
| **BMI, mean (SD), kg/m<sup>2</sup>** | 27.4 (4.8) | 25.9 (4.4) | 28.9 (3.8) | 26.9 (4.3) | 28.9 (4.5) | 26.9 (4.5) | 28.8 (4.8) | 29.3 (4.1) |

**Figure 1.**
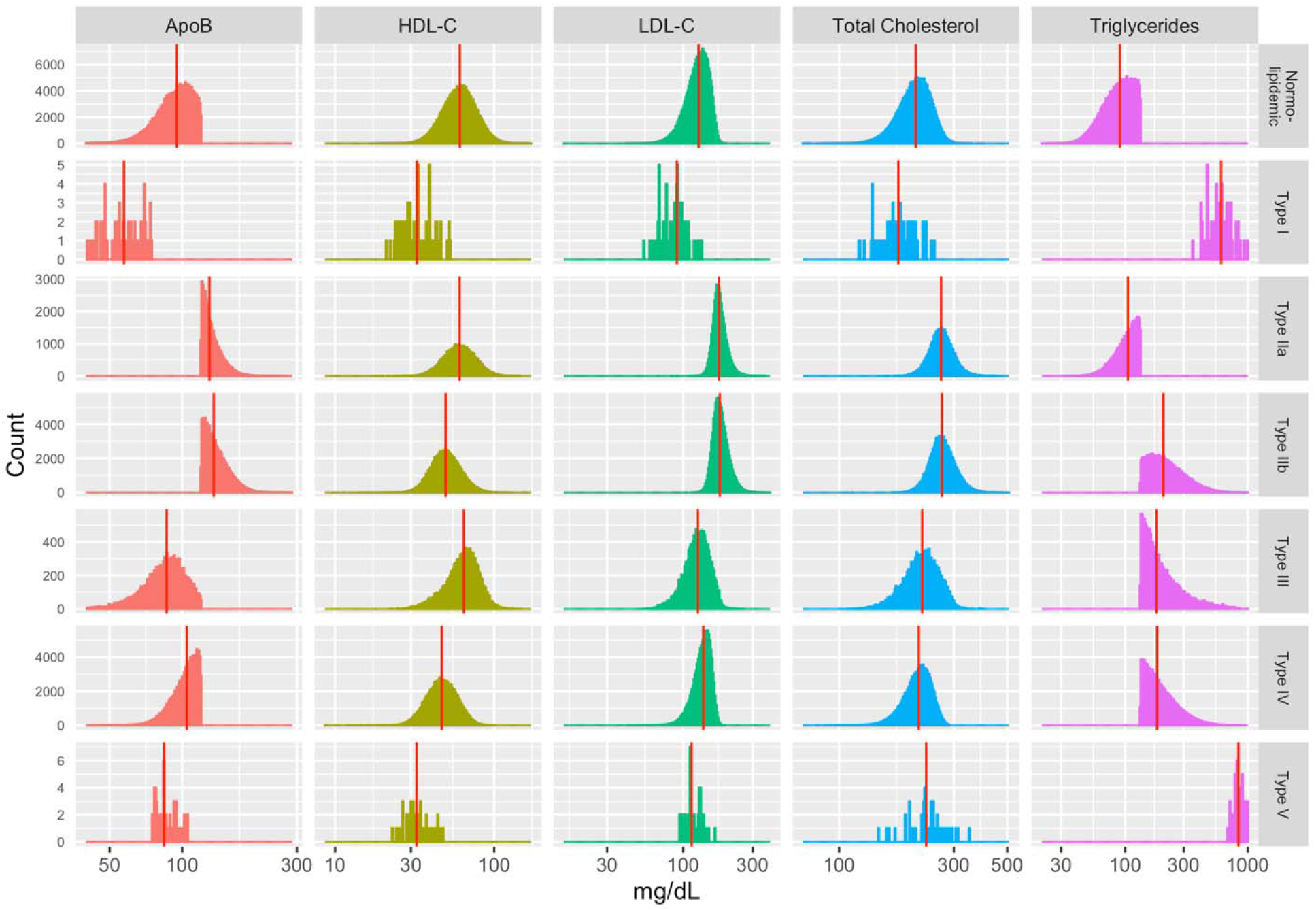
Distribution of lipids among normolipidemics and FLL classes. Lipids are presented as mg/dL on the log(10) scale, with vertical dash marking distribution median. Abbreviations: ‘ApoB’ = apolipoprotein B; ‘HDL-C’ = high-density lipoprotein cholesterol; ‘LDL-C’ = directly measured low-density lipoprotein cholesterol. ApoB, LDL-C, and total cholesterol are corrected for baseline statin prescription, as outlined in *Methods*.

### Risk of incident CAD by FLL dyslipoproteinemia class

Over median 11.1 years of follow-up, adjusted for age, sex, smoking status, BMI, hypertension, type 2 diabetes mellitus, statin prescription, genotyping array, and the first five principal components of ancestry, participants with FLL type IIb had the highest risk of incident CAD (HR 1.92, 95% CI 1.84-2.01, P<0.001) compared to normolipidemics, followed by type IIa (1.51, 95% CI 1.42-1.61, P<0.001) and type IV (HR 1.28, 95% CI 1.22-1.34, P<0.001) (**Figure 2, Supplementary Table 2**). Type III individuals were not at statistically increased risk relative to normolipidemics (HR 1.05, 95% CI 0.93-1.18, P=0.44). Adjusting the model further for either apoB or non-HDL-C attenuates the effect of FLL class on incident CAD, with preservation of increased hazard among individuals with the type IIb phenotype compared to other common FLL classes (**Supplementary Tables 3 and 4** for models additionally adjusting for apoB and non-HDL-C, respectively). Type III reaches nominal significance only in the model adjusted for apoB (HR 1.08, 95% CI 1.02-1.29, P=0.02). When patients prescribed lipid-lowering medications at baseline were excluded from the primary survival analysis, a similar pattern was observed of increased hazard for type IIb versus the other FLL disorders against normolipidemics (**Supplementary Table 5**).

**Figure 2.**
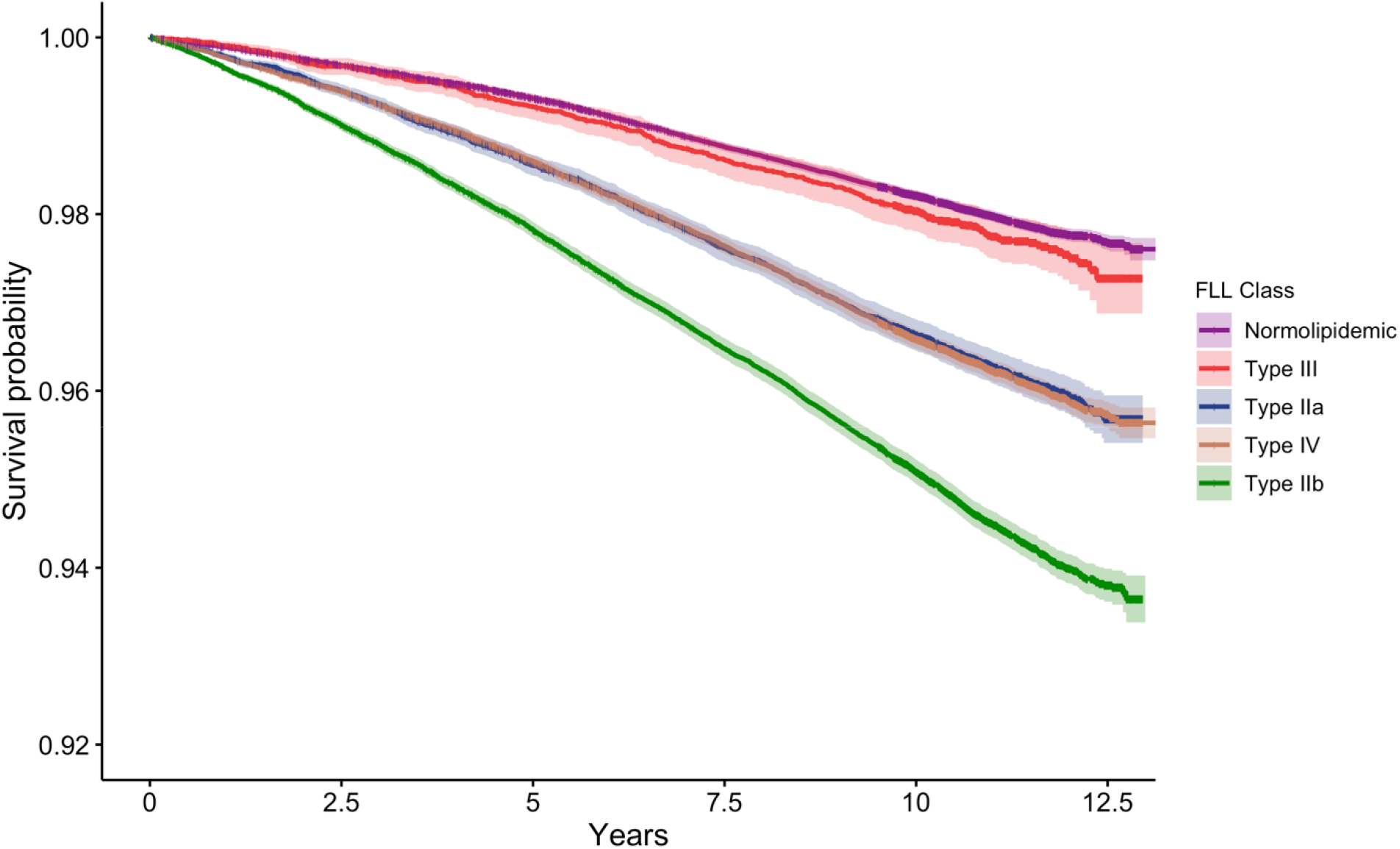
Survival free from incident CAD according to FLL class. Individuals with FLL dyslipoproteinemias were followed for a median of 11.1 years for incident CAD. Abbreviations: ‘CAD’ = coronary artery disease; ‘FLL’ = Fredrickson-Levy-Lees; ‘Normolipidemic’ = not meeting criteria for any FLL disorder.

Across strata of non-HDL-C (130-160, 160-190, 190-220, and 220-250 mg/dL), the type IIb pattern was associated with increased risk of incident CAD compared to types IIa, III, and IV when compared to stratum-matched normolipidemic individuals. In random-effects meta-analysis across the non-HDL-C strata, type IIb conferred a 1.45-fold independent hazard (95% CI 1.30-1.60) for incident CAD (Figure 3). Risk was elevated for types IV and IIa (HR 1.18, 95% CI 1.10-1.30 and HR 1.17, 95% CI 1.04-1.30, respectively) but not for type III individuals (**Supplementary Figure 2**).

**Figure 3.**
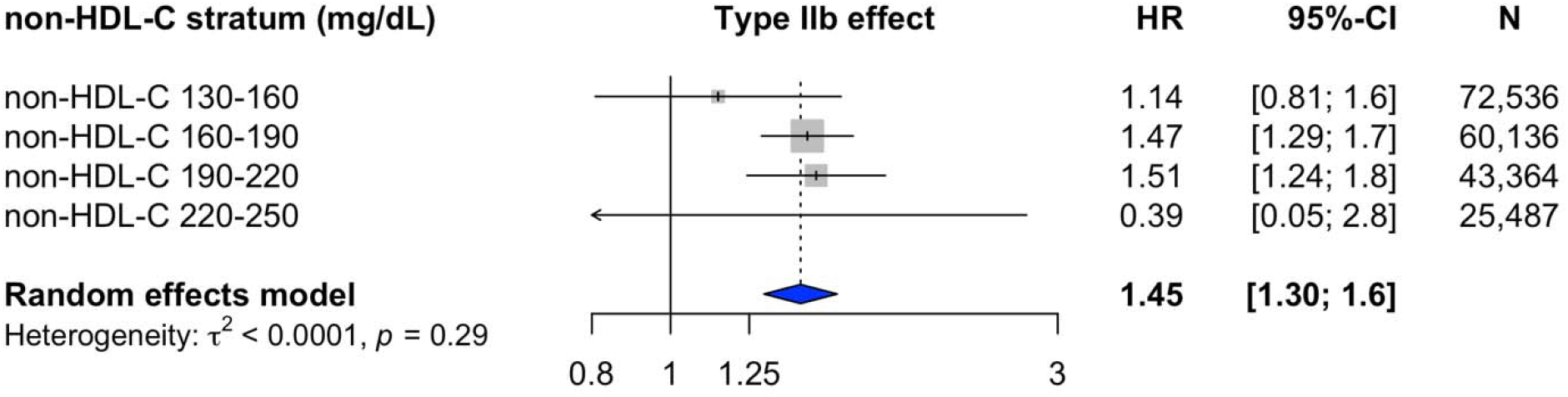
FLL type IIb confers elevated CAD risk across non-HDL-C strata. Meta-analysis of FLL type IIb effect compared to normolipidemic individuals across non-HDL-C strata. Individuals meeting criteria for FLL type IIb were compared against stratum-matched normolipidemic individuals across four strata of non-HDL-C. Random-effects meta-analysis reported overall type IIb effect. Abbreviations: ‘Non-HDL-C’ = non-high-density lipoprotein cholesterol; ‘HR’ = hazard ratio; ‘CI’ = confidence interval; ‘N’=total individuals in a given stratum; *τ*^2^ = effect size variance estimated by restricted maximum likelihood method.

### Phenome-wide association testing in UK Biobank

Unbiased phenome-wide association testing was performed to screen for associations between FLL classes and combined prevalent or incident phenotypes among UKBB participants, adjusted for age, sex, genotyping array, and the first five principal components of ancestry. A total of 1584 phenotypes were considered in the UKBB. Effect estimates (odds ratio) between FLL type and phecode for the union of the top 50 associations for each FLL type are shown in **Supplementary Figure 3**. Representative top associations across the four phenotypes are shown in **Figure 4**. While type IIa was strongly associated with atherosclerotic cardiovascular disease (ASCVD) phenotypes, effects were greater for type IIb, which was also accompanied by strong associations with diabetes mellitus, liver disease, and obesity. Type IV was also similarly associated with diabetes mellitus, liver disease, and obesity, but effects for ASCVD were less. Type III was most strongly associated with alcohol-related disorders and malnutrition along with liver disease but without significant associations for diabetes or CAD.

**Figure 4.**
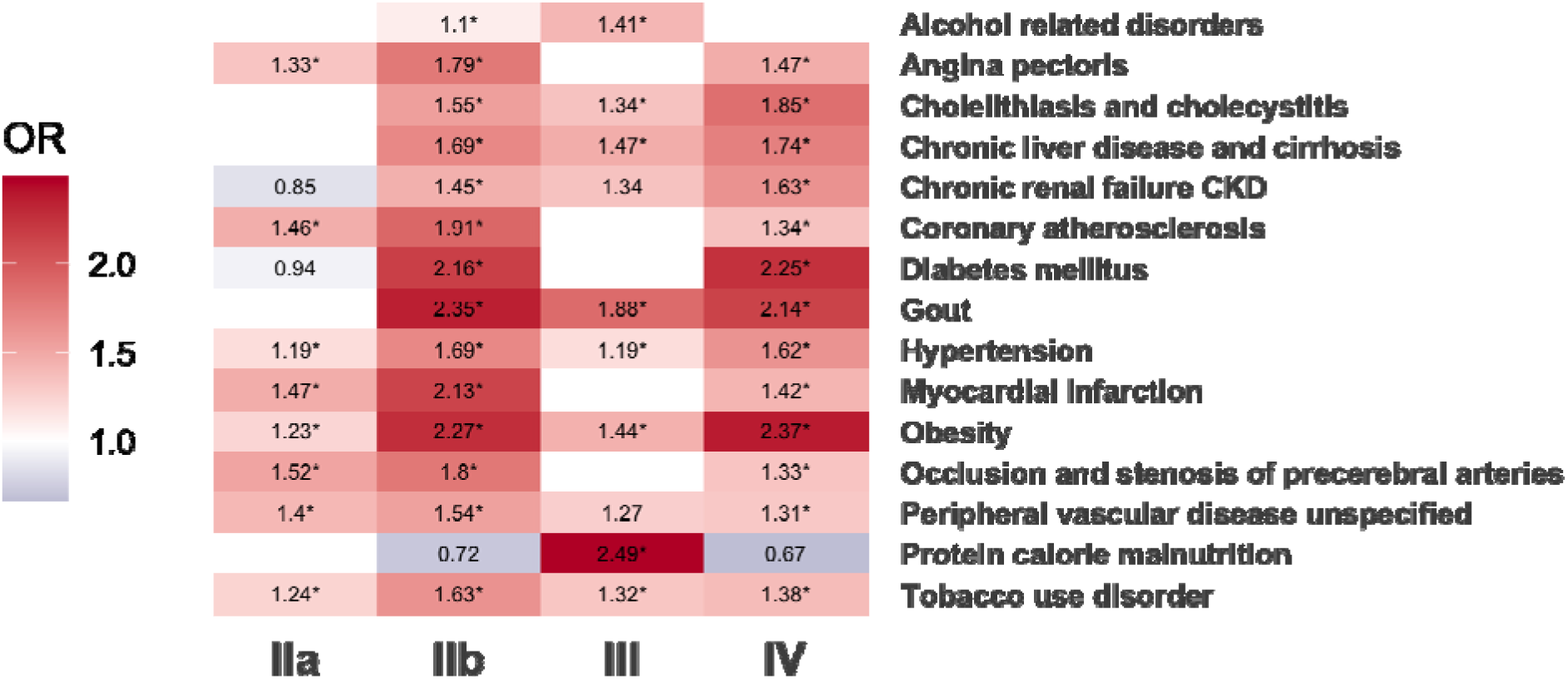
Phenome-wide association of lipid patterns yields distinct phenotypic patterns. PheWAS UKBB of representative phenotypes reaching phenome-wide significance (Bonferroni adjusted P<3.2×10^−5^) relative to normolipidemics. OR is presented only when nominal P value of P<0.05 is reached and are marked with “*” if reaching phenome-wide significance. Blank cells reflect exposure-phenotype associations P>0.05.

### Genomic architecture of FLL dyslipoproteinemias

Among 98,587 individuals with any FLL dyslipoproteinemia with WES, only 2,020 (2.05%) carried a disruptive variant in a monogenic dyslipidemia gene. Type IIa had the greatest prevalence (479 of 15,309 [3.1%]) of a disruptive monogenic dyslipidemia gene, most commonly in *LDLR* (**Supplementary Table 6, Supplementary Figure 4**). We also assessed *APOE* genotype status among study participants. Notably, the *APOE*-2/2 genotype, which is classically associated with remnant hypercholesterolemia, was markedly enriched among FLL type III individuals (11.4% prevalence) relative to the other FLL phenotypes and was rare among normolipidemics (0.5% prevalence) (**Supplementary Table 7, Supplementary Figure 5**).

Common variants were assessed for relevance separately for FLL types IIa, IIb, III, and IV (types I and V were excluded due to low case counts) against normolipidemic individuals through GWAS. Among 418603 unrelated individuals included, there were 57 with type I, 37,211 with type IIa, 88,007 with type IIb, 13,030 with type III, 102,677 with type IV, 45 with type V, and 177,576 normolipidemics. These independent GWAS identify 77, 210, 31, and 99 variants contributing to FLL types IIa, IIb, III, and IV, respectively (**Supplementary Tables 8-11**). Taken together, these represent a total of 250 unique genome-wide significant (P < 5×10^−8^) loci contributing to any FLL class (**Figure 5, Supplementary Table 12**). Of the total associated loci, 13 were shared across all phenotypes (**Supplementary Table 13**), all of which are previously described lipid-associated genes. Unique loci for types IIa, IIb, III, and IV, number 12, 97, 5, and 21, respectively (**Supplementary Figure 7, Supplementary Table 14)**.

**Figure 5.**
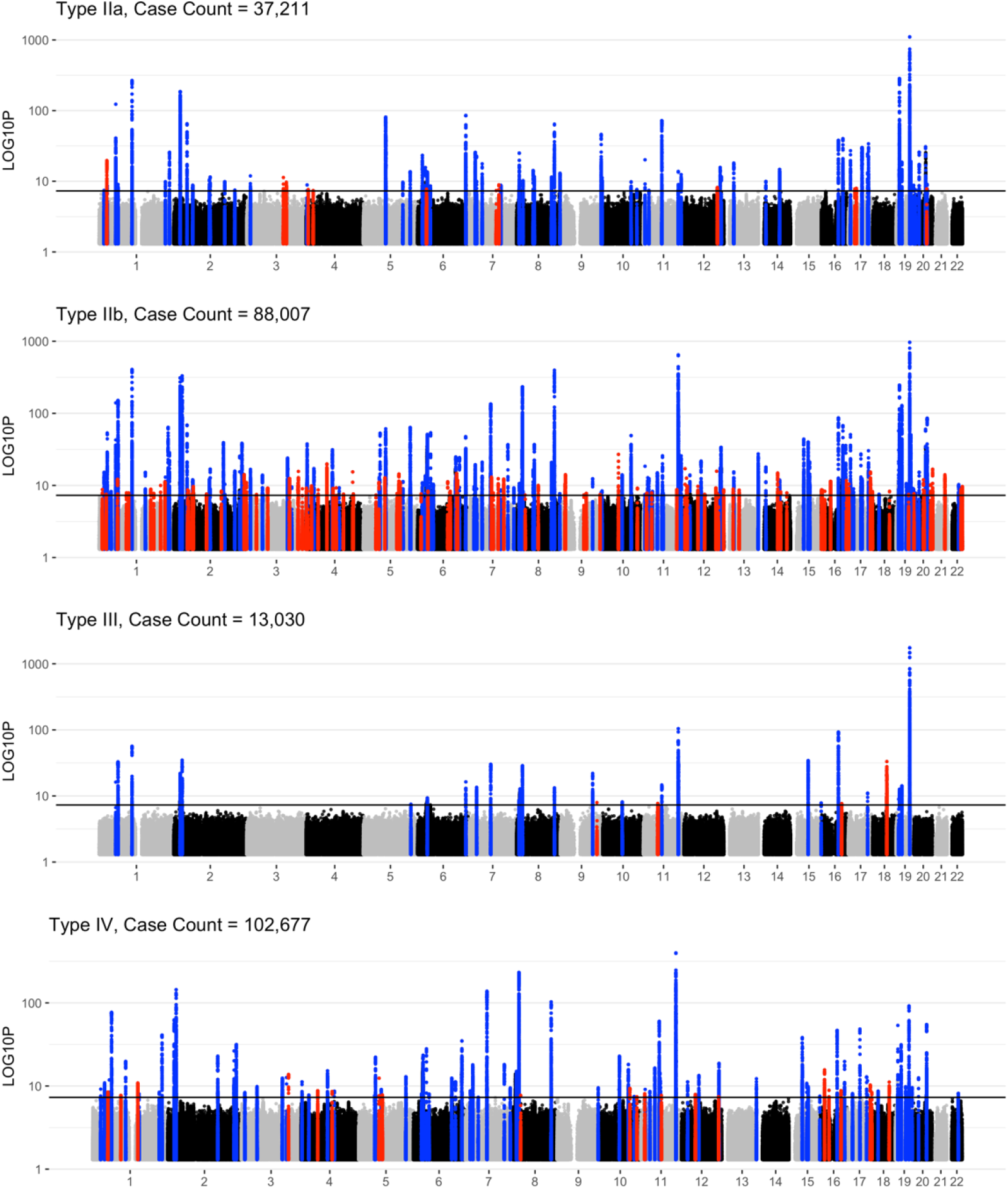
Shared and distinct genetic loci associated with FLL classes. Independent GWAS were performed using REGENIE software for each of the common FLL phenotypes (IIa, IIb, III, and IV) against normolipidemic individuals. Independent genome-wide significant SNPs define +/− 500kbp loci, which are highlighted in red or blue. Red loci are unique to the phenotype, and blue loci are shared between two or more phenotypes. X-axis reflects genomic location labeled according to chromosome 1-22. Y-axis displays −log_10_(P-value) on the log_10_ scale.

When compared to a recent GWAS of similar size in the Million Veterans Program for individual lipid concentrations (inclusive of HDL-C, LDL-C, TC, and TG), several loci (72 of 250 [28.8%]) are novel(38). When further compared against published studies in the GWAS Catalog(39), 14 of the total loci are not reported to be associated with any of HDL-C, LDL-C, TG, total-C, or apoB concentrations (**Supplementary Table 15**). Examples include rs17576576, which associates with the FLL IIa phenotype and is upstream of peroxisome proliferator-activated receptor gamma, coactivator 1 alpha (*PPARGC1A*), and rs3856522, which associates with the FLL IIb phenotype and is an exonic variant of ATPase homolog 2 pseudogene (*AHSA2P*). Rs141955254 is the sole novel purported association with the Type III phenotype and is annotated as an intergenic variant in proximity to BMP/retinoic acid inducible neural specific 1 (*BRINP1*).

Independent variants reported in the recent CARDIoGRAM-C4D Consortium CAD GWAS meta-analysis were extracted from FLL GWAS summary statistics for each phenotype(40). There was substantial overlap between CAD and FLL phenotype variants. Of the genome-wide significant CAD variants extracted from FLL GWAS summary statistics, 42.8% of variants were nominally significant at P<0.05 for type IIb (16.9% genome-wide), 32.2% for type IIa (12.7%), 27.1% for type IV (11.4%), and 24.2% for type III (6.4%) (**Supplementary Figure 8**). Direction of effect was generally shared between CAD variants and FLL variants for phenotypes IIa and IIb but not for phenotypes III and IV (**Supplementary Figure 9**). For example, *APOB* locus variants increasing the risk of CAD increased the risk of FLL types IIa and IIb but decreased the risk of phenotypes III and IV. Further, we observe that certain loci strongly affect risk of one condition while only modestly affecting the comparator phenotype. *LPA* variants have a strong influence on CAD risk but more modest influence on the FLL phenotypes.

Polygenic priority scoring ranked protein-coding genes from the common variants GWAS results for FLL types IIa, IIb, III, and IV using external multi-omics data. **Supplementary Figure 10** shows the union of the top 20 genetic contributors to each phenotype according to PoPS score. *APOE* was the top-ranked gene across all phenotypes, and common variants linked to Mendelian lipid-related genes were prioritized for all phenotypes, including *LDLR, APOB, SCARB1*, and the *APOA5/A4/C3/A1* locus. *LPL* was prioritized more highly for the hypertriglyceridemic types (i.e., IIb, III, and IV) than for type IIa. Some prioritized novel genes are platelet derived growth factor subunit B (*PDGFB*) for type IV, BRCA1 DNA repair associated (*BRCA1*) and beta-1,4-galactosyltransferase 1 (*B4GALT1*) for type III, and insulin receptor substrate 1 (*IRS1*) and insulin like growth factor 1 (*IGF1*) for type IV.

LD score regression estimated heritabilities and genetic correlations among the FLL dyslipoproteinemias (**Supplementary Figure 11**). Observed SNP heritabilities (standard errors [SE]) for FLL types IIa, IIb, III, and IV were 0.37 (0.1), 0.54 (0.09), 0.23 (0.29), and 0.28 (0.04), respectively. Pairwise genetic correlations were greatest between FLL types IV and III (0.96, SE 0.03), types IIb and III (0.92, SE 0.05), and types IIb and IV (0.82, SE 0.02). The least genetic correlations were observed between types IIa and III (0.02, SE 0.05) and IIa and IV (0.09, SE 0.01).

### Sensitivity analysis: Accounting for fasting time

A minority of individuals (N=18488) fasted for greater than or equal to 8 hours prior to baseline blood draw. Utilizing the same algorithm for FLL classification, a similar prevalence of dyslipoproteinemia was observed: 52.3% were dyslipoproteinemic among fasting individuals compared to 57.5% among all individuals. No significant different in the prevalences of FLL types I, IIb, III, and V was noted, whereas types IIa and IV were less common among fasting individuals (**Supplementary Table 17**).

Repeating GWAS with fasting time as a covariate, restricted to top independent SNPs for each phenotype, yielded no significant different in effect estimates or association P-value for any of the FLL disorders (**Supplementary Figure 12**).

## Discussion

In a large contemporary prospective cohort, we identified individuals with one of six FLL dyslipoproteinemias using an apoB-based classification algorithm. The prevalence of FLL disorders was high, affecting more than half of individuals. In both primary analysis and sensitivity analyses controlling for apoB or non-HDL-C, the type IIb phenotype conferred the highest risk of incident CAD compared to the remaining FLL dyslipoproteinemias; importantly, risk was also elevated for types IIa and IV in all analyses. FLL patterns exhibited diverse phenotypic relationships, including CAD for types IIa, IIb, and IV; diabetes and liver disease for types IIb and IV; and alcohol-related disorders and liver disease for type III. Common variant analysis using GWAS for the four prevalent FLL disorders revealed multiple genetic contributions to each, with novel loci detected not previously associated with individual lipids. Monogenic dyslipidemia variants were most common among type IIa individuals, usually affecting *LDLR*, but were rare overall (i.e., 2% prevalence), and heritability analyses highlighted the polygenicity of the FLL phenotypes.

Our study adds to the growing evidence that FLL dyslipoproteinemias are underrecognized in contemporary Western populations(3,4). We show that identification of combinations of lipid derangement – as initially conceived by Fredrickson and colleagues – allows for refinement of CAD risk, as seen with the type IIb pattern when traditional lipid parameters are controlled for. Characterizing combinations of lipids furthermore yields distinct patterns of cardiometabolic risk not captured by single lipid parameters. While both types IIa and IIb are associated with CAD risk, only the latter carries significant risk independent of apoB concentration and also carries risk for diabetes and liver disease. While current guidelines prioritize FH, a pattern consistent with type IIa, for early preventive therapies, our observations support extension to type IIb alongside strategies to prevent diabetes onset.

The FLL type III phenotype was not associated with CAD in our study, in contrast to prior descriptions. As previously reported, the apoB-based scheme may be more sensitive and less specific for severe versions of type III dysbetalipoproteinemia in comparison to ultracentrifugation(4,41). Further, given that type III is reported to be a labile finding even among *APOE*-2/2 carriers and likely highly influenced by environmental exposures, follow-up time in our model may not accurately reflect total remnant cholesterol exposure. Finally, the subclinical atherosclerosis described in contemporary cohorts with FLL type III individuals may not manifest as clinical ASCVD in the relatively young and healthy UKBB cohort.

Our genetic studies and downstream analysis revealed several interesting findings. First, canonical lipid loci are important contributors to each of the four most common FLL phenotypes, but non-lipid loci appear to be associated as well. For example, we identify the *IRS1* locus as an important contributor to both type IIb and IV phenotypes. A variant ~500kb upstream of *IRS1* (rs2943641) is previously reported to be associated with a 35% increased risk of type 2 diabetes, the pathophysiologic basis of which appears to be impaired insulin sensitivity (and thus increased serum insulin) rather than altered beta cell homeostasis(42). This variant is strongly associated with both type IIb and IV phenotypes and is in close proximity and LD with lead CAD risk variant rs2943634(43). Thus, our strategy of combining multiple lipid derangements allows us to link insulin resistance, lipid abnormalities, and CAD risk for a variant likely driving CAD risk via a mechanism distinct from hypercholesterolemia. Second, while there may be important instances of monogenic inheritance for each of the FLL types, polygenicity is a far more common pattern of inheritance. This is consistent with prior genetic studies of individuals with severe hypercholesterolemia (akin to FLL type IIa) and with combined hyperlipidemia (akin to FLL type IIb)(13). In the case of the type IIb phenotype, our demonstration of high concordance between CAD risk loci and type IIb risk loci is consistent with the finding of high atherogenicity. Further, we find that, though concentrated among type III individuals, the *APOE*-2/2 genotype is not necessary for its expression; rather, polygenic susceptibility appears sufficient. Third, SNP heritability is high, and genetic correlation analysis reveals clusters of similarity – the hypertriglyceridemic conditions (types IIb, III, and IV) share greater genetic overlap than either types IIa and IIb or IIa with the other phenotypes.

Phenotype association testing reveals both previously reported and novel disease associations. As expected, type IIa was associated with traditional ASCVD phenotypes, and hypertriglyceridemic types IIb and IV were associated with obesity and diabetes mellitus, with type IV being associated with more biliary disorders and less strongly for ASCVD, suggesting that these may reflect a spectrum of a similar dyslipoproteinemia associated with metabolic syndrome. Interestingly, FLL type III was associated with alcohol-related disorders and both malnutrition and hyperalimentation. This finding suggests that the type III pattern may be related to an exposure not well captured by these descriptors (e.g., liver disease) or simply cannot be explained by a single genetic by exposure axis.

Combining our genetic and phenotypic findings reveals patterns of association for specific FLL classes and highlights opportunities for precision medicine approaches for dyslipidemia treatment. For example, we find that variants at *RBM47* are uniquely associated with the type IIb phenotype. The *RBM47* locus is recently reported as a putative novel CAD risk locus among individuals of Middle Eastern ancestry(44). Further, in exome sequencing of diverse ancestry populations there was an association between the burden of deleterious variants in *RBM47* and TG/HDL-C ratio, and specific variants have been shown to significantly alter apoC-III concentrations(45). This may indicate that the type IIb phenotype would benefit from apo-CIII-lowering therapies in addition to LDL-C-lowering therapies. We also found the *FAM13A* locus to be uniquely associated with the type IIb phenotype (lead variant rs6824451, OR 1.04, *P*=7.84×10^−10^). This variant also increased risk for the type IV phenotype, although it did not reach genome-wide significance (OR 1.03, P=7.48×10^−7^). *FAM13A* encodes family with sequence similarity 13 member A, which has been implicated in body fat distribution in humans and visceral to subcutaneous adipose tissue ratio in mice via effects on adipocyte differentiation(46); a functional genomic study in adipocytes prioritized *FAM13A* as a causal gene in insulin resistance(47). Overall, these findings point to a common mechanism connecting abnormal adipocyte homeostasis with lipid phenotype among hypertriglyceridemic FLL types IIb and IV.

This study has several limitations. First, the UKBB is a predominantly European ancestry cohort, and studies in ancestrally diverse populations are necessary to test whether these findings generalize to non-European groups. Second, we relied on non-fasting lipids (the UKBB Biobank standard) to assign Fredrickson class. Reassuringly, we found that when we limited the population to those with >= 8 hour fast prior to the baseline lab draw, a similar proportion of participants met criteria for a FLL disorder, suggesting this is simply a high prevalence population. Further evidence of high baseline dyslipoproteinemia is that type IIa prevalence is double that in other contemporary cohorts (8.9% versus 4-5%), despite apoB not being significantly influenced by the post-prandial state(22,48). Importantly, though isolated hypertriglyceridemia (FLL type IV) is classically defined in the fasting state, post-prandial hypertriglyceridemia has equal implications for cardiovascular risk (49). Our approach of applying the FLL framework to non-fasting samples permits clinical and genetic discovery on a scale not feasible when restricting to fasting-only samples, and we show that genetic results are unaffected by fasting time.

By accounting for patterns of change in multiple lipoprotein subfractions, the FLL classification scheme describes an underlying pathophysiologic disturbance with specificity that individual lipoprotein fractions cannot. Individuals with the type IIb pattern are at elevated risk of ASCVD relative to other dyslipoproteinemic individuals at equivalent non-HDL-C and thus merit intensive risk reduction and attention to the associated disease states that may contribute to the phenotype. Differential genetic risk factors underlying the FLL types highlights opportunities for precision medicine strategies aimed at lipid patterns beyond LDL-C alone.

## Data Availability

The data underlying this article were accessed from the UK Biobank under Application number 7089. UK Biobank individual-level data are available for request by application (www.ukbiobank.ac.uk).

## Funding

Dr. Gilliland is partially supported by the National Heart, Lung, and Blood Institute T32 grant 5T32HL125232. Dr. Dron is supported by the Canadian Institute of Health Research as a Banting Postdoctoral Research Fellow. Drs. Peloso and Natarajan are supported by grants from the National Heart, Lung, and Blood Institute (R01HL142711, R01HL127564). In addition, Dr. Natarajan is supported by grants from the National Heart Lung and Blood Institute (R01HL148050, R01HL151283, R01HL148565, R01HL135242, R01HL151152), National Institute of Diabetes and Digestive and Kidney Diseases (R01DK125782), Fondation Leducq (TNE-18CVD04), and Massachusetts General Hospital (Paul and Phyllis Fireman Endowed Chair in Vascular Medicine).

## Acknowledgements

The authors would like to acknowledge and thank the staff, investigators, and participants of the UK Biobank.

## Disclosures

Dr. Natarajan reports grant support from Amgen, Apple, AstraZeneca, Boston Scientific, and Novartis, spousal employment and equity at Vertex, consulting income from Apple, AstraZeneca, Novartis, Genentech / Roche, Blackstone Life Sciences, Foresite Labs, and TenSixteen Bio, and is a scientific advisor board member and shareholder of TenSixteen Bio and geneXwell, all unrelated to this work. The other authors report no disclosures.

## Notes

### Author Declarations

This research was conducted under UK Biobank application number 7089. The Mass General Brigham Institutional Review Board approved secondary use of these data (2013P001840).

### Summary of Updates

Supplemental figures added

