## Supplementary figures for "Genetic Architecture And Clinical Outcomes Of The Fredrickson-Levy-Lees Dyslipoproteinemias"

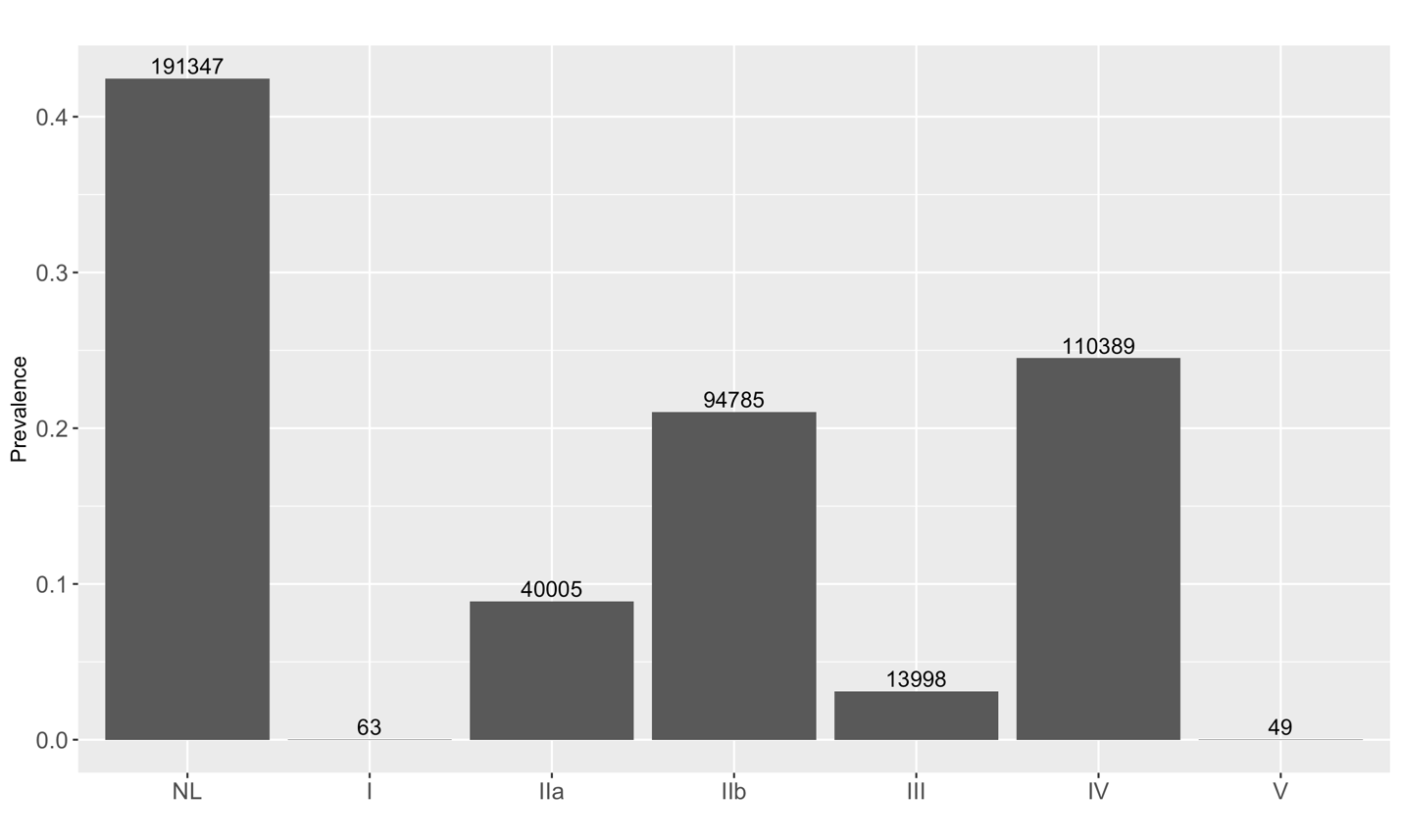

**Supplementary Figure 1. FLL disorders are highly prevalent among UKBB participants.** Prevalence of FLL disorders within UKBB. Bars represent percentage of study participants meeting criteria for each FLL dyslipoproteinemia. Counts are presented on top of each bar. Y-axis reflects percentage of total (N_total_=450636). ‘NL’ = normolipidemic; ‘I’ = FLL type I; ‘IIa’ = FLL type IIa; ‘IIb’ = FLL type IIb; ‘III’ = FLL type III; ‘IV’ = FLL type IV; and ‘V’ = FLL type V.

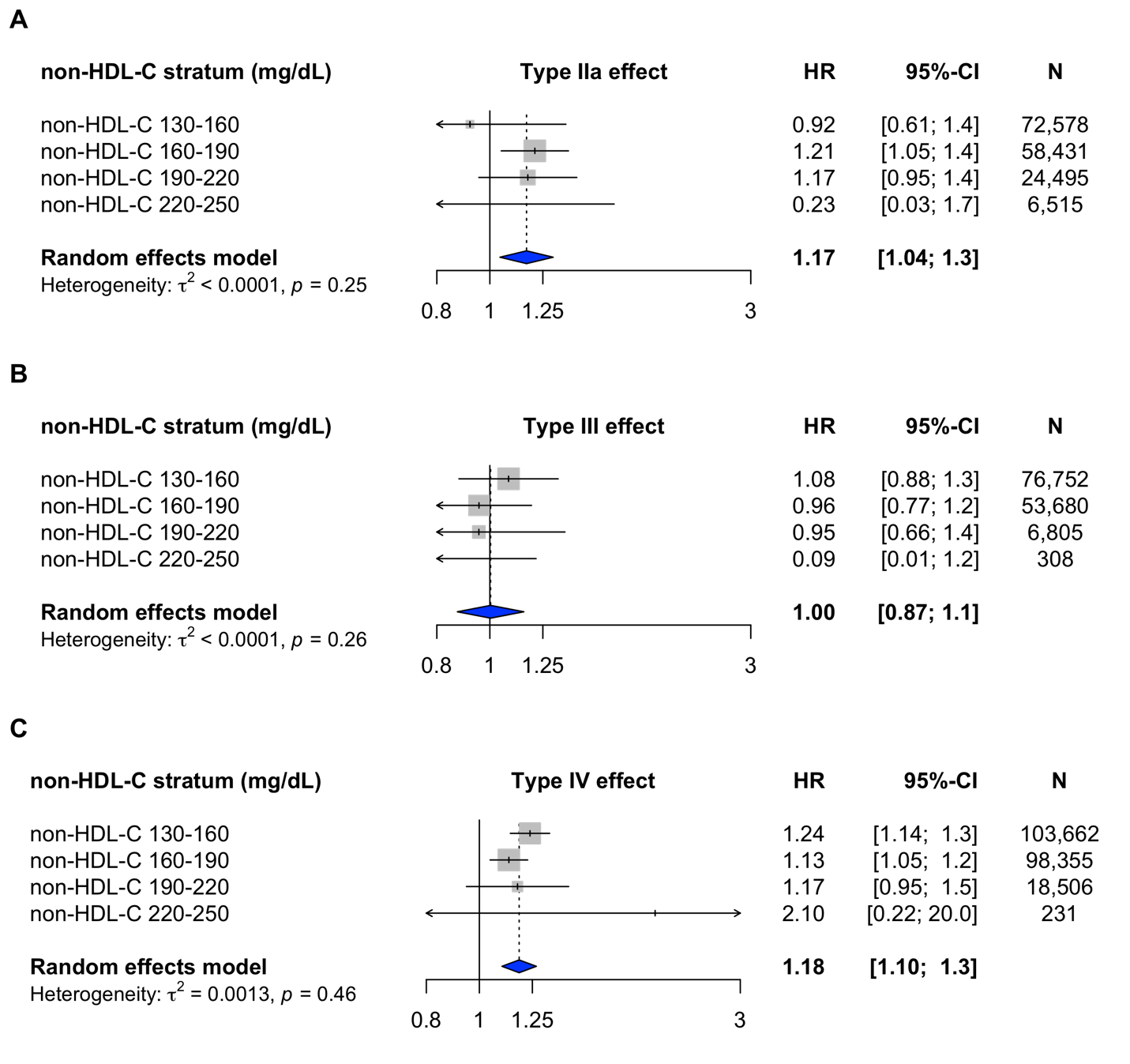

**Supplementary Figure 2. FLL types IIa and IV, but not type FLL type III, confers elevated CAD risk across non-HDL-C strata.** Meta-analysis of FLL types IIa, III, and IV effect compared to normolipidemic individuals across non-HDL-C strata. Individuals meeting criteria for FLL type IIa (A), FLL type III (B), and FLL type IV (C) were compared against stratum-matched normolipidemic individuals across four strata of non-HDL-C. Random-effects meta-analysis reported overall FLL class effect. Abbreviations: ‘non-HDL-C’ = non-high-density lipoprotein cholesterol; ‘HR’ = hazard ratio; ‘CI’ = confidence interval; ‘N’=total individuals in a given stratum; τ^2^ = effect size variance estimated by restricted maximum likelihood method.
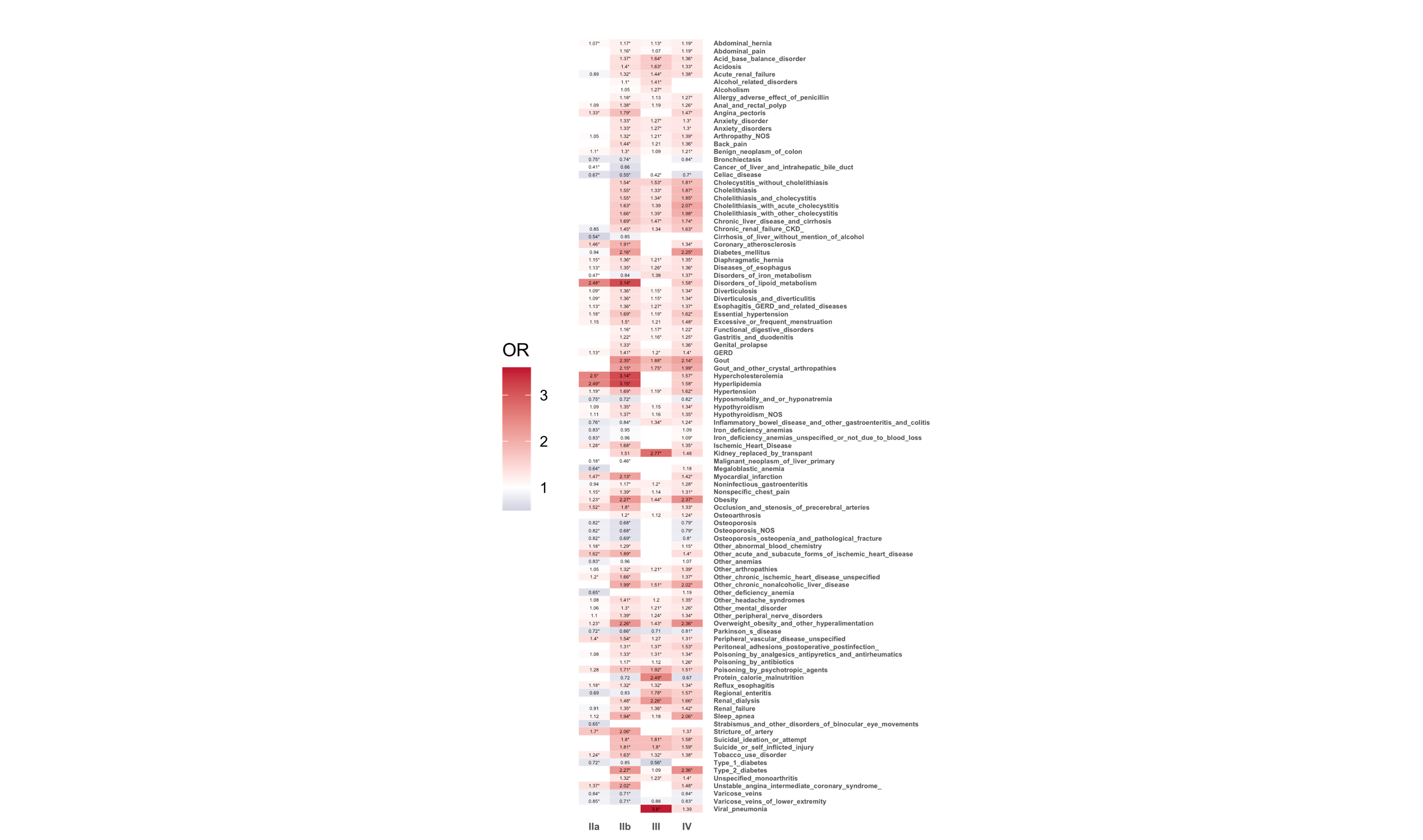

**Supplementary Figure 3. Top associations between FLL disorders and UKBB phenotypes.** Union of the top 50 associations between FLL phenotypes IIa, IIb, III, and IV and curated phecodes (see *Methods* for details), yielding a total of 100 top associations. Shading reflects odds ratio of association, with red shades positively associated and blue shaded negatively associated. OR is presented only when nominal P value of P<0.05 is reached and are marked with “*” if reaching phenome-wide significance (P <3.3x10^-5^). Blank cells reflect exposure-phenotype association P>0.05.

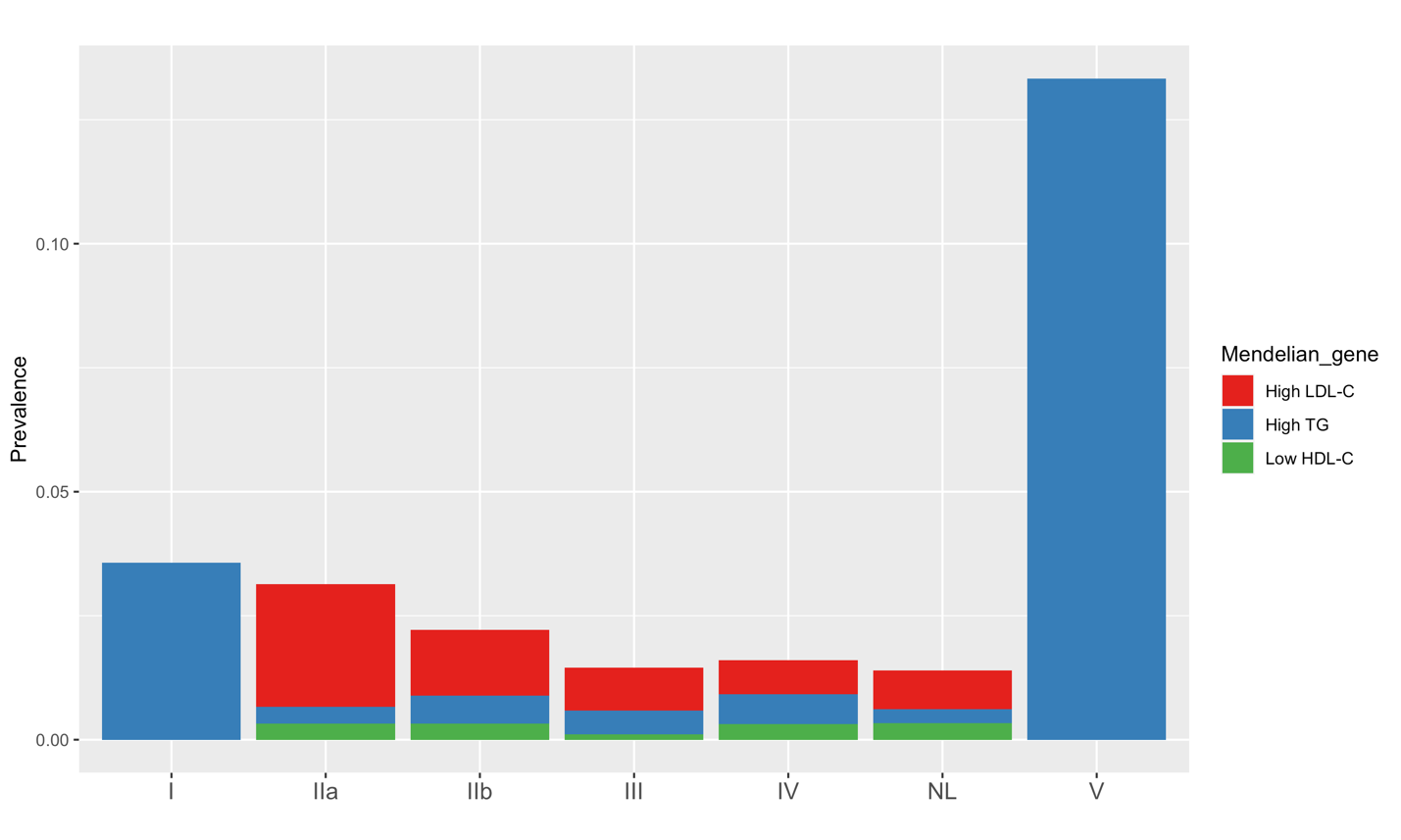

**Supplementary Figure 4. Monogenic dyslipidemia mutations are rare among UKBB participants.** Individuals included in the genome-wide association study of FLL phenotypes were queried for monogenic dyslipidemia mutations associated with high LDL-C (*LDLR*, *APOB*, *PCSK9*, *ABCG5*, *ABCG8*), high TG (*APOA5*, *APOE*, *LPL*, *APOC2*, *GPIHBP1*, *LMF1*), and low HDL-C (*ABCA1*, *APOA1*, *LCAT*). X-axis is FLL phenotype (‘NL’ = normolipidemic), and Y-axis is population prevalence of monogenic dyslipidemia mutations.

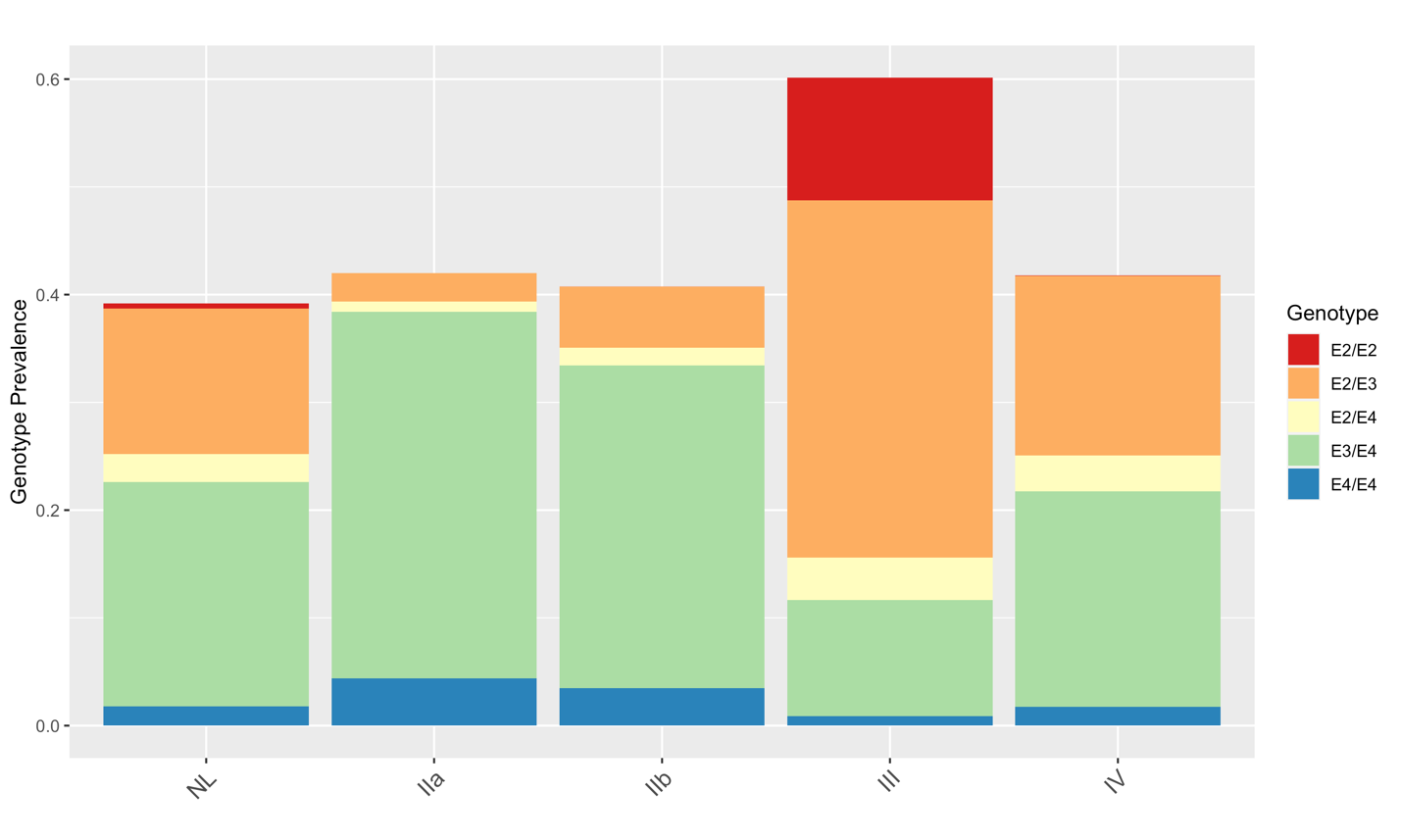

**Supplementary Figure 5. *APOE*-2/2 genotype is enriched among UKBB participants with the type III phenotype.** Bar plots reflect distribution of *APOE*-WT (i.e., *APOE*-3/3) genotypes among UKBB participants according to FLL phenotype. FLL types I and V are not included due to low case counts. ‘NL’ = normolipidemic; ‘IIa’ = FLL type IIa; ‘IIb’ = FLL type IIb; ‘III’ = FLL type III; ‘IV’ = FLL type IV.

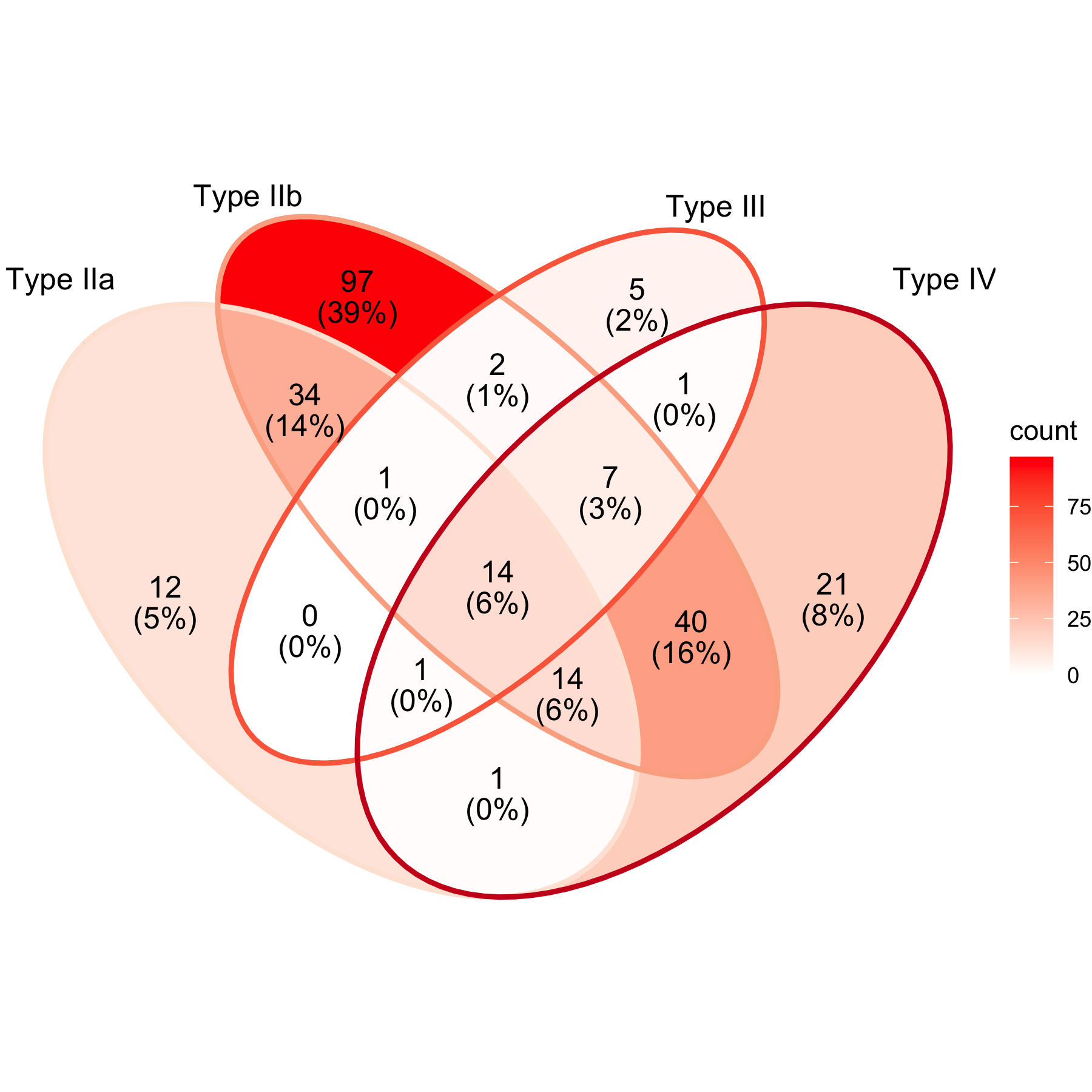

**Supplementary Figure 6. Loci overlap between FLL classes.** Overlap of 250 total genetic loci associated with FLL types IIa, IIb, III, and IV. Presented as loci counts (% of total loci).

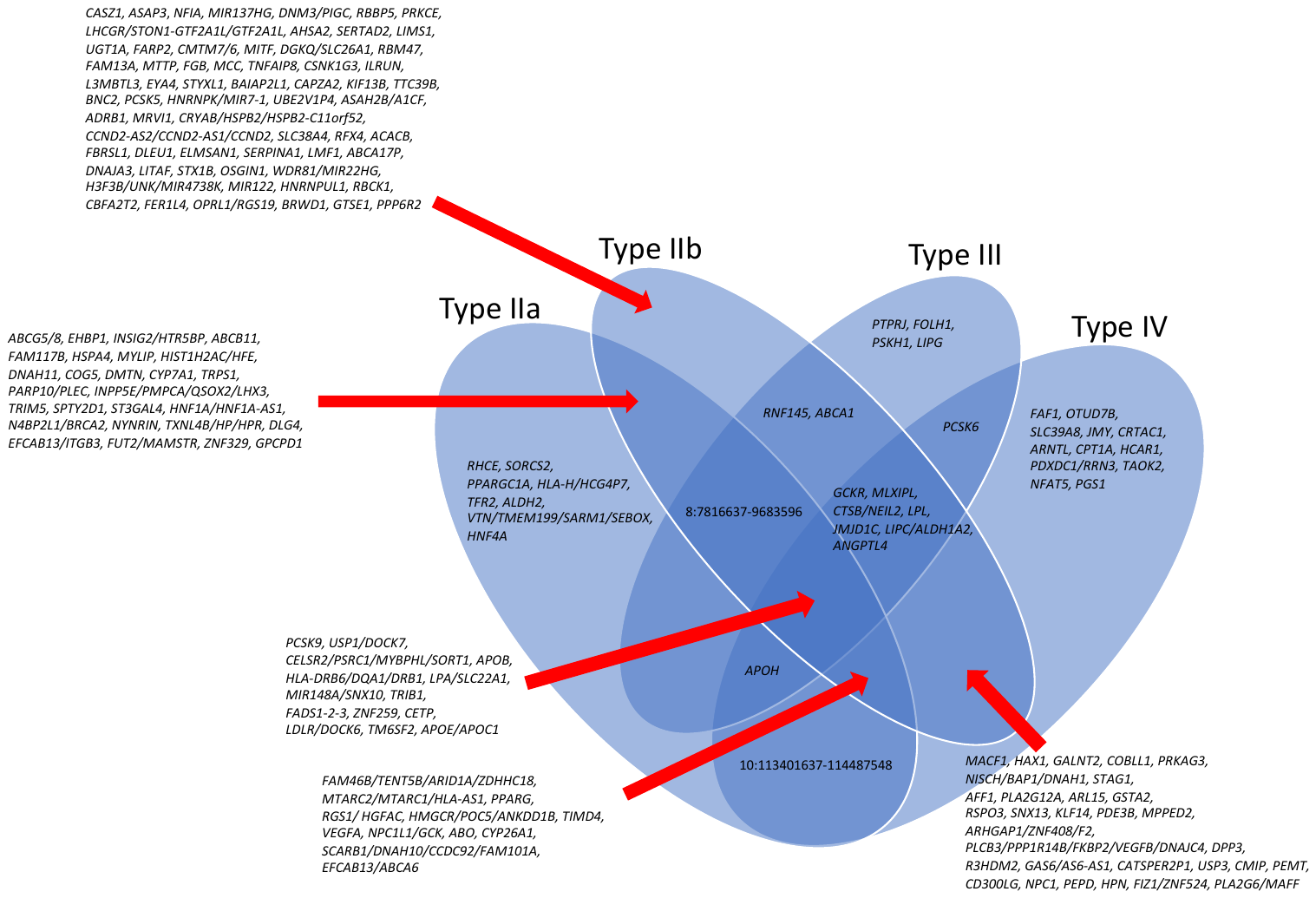

**Supplementary Figure 7. Loci overlap between FLL classes, genes included.** Overlap of genetic loci associated with FLL types IIa, IIb, III, and IV. Loci are included when Ensembl Variant Effect Predictor (<https://useast.ensembl.org/info/docs/tools/vep/index.html>) annotated a protein-coding gene. There are no genetic loci overlapping between FLL Types IIa and III.

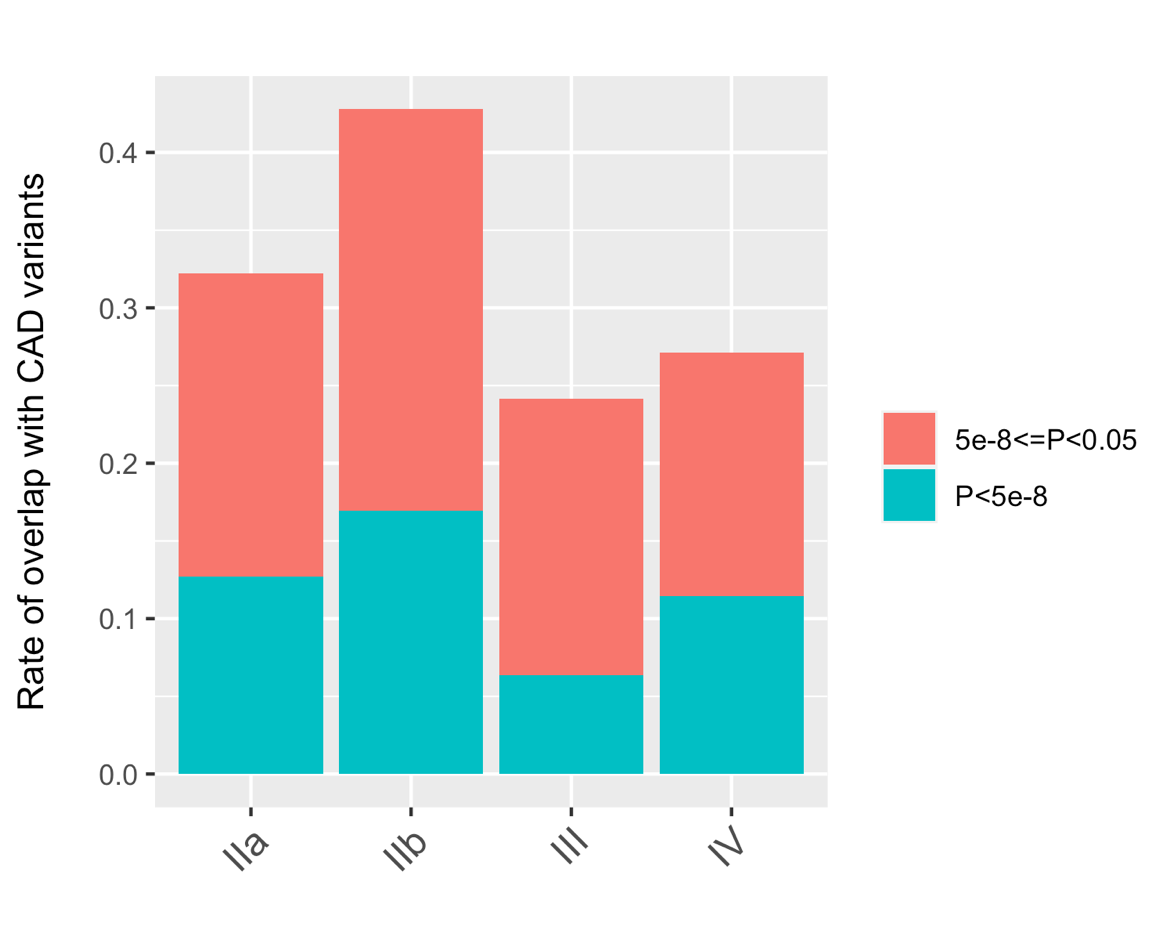

**Supplementary Figure 8. Rate of overlap of FLL SNPs previously reported as top independent signals in CAD GWAS.** Of 241 top independent SNPs reported in CAD GWAS, 237 were available in the FLL GWAS. For each of FLL phenotypes IIa, IIb, III, IV, top CAD loci are reported if they are associated with a given FLL phenotype at P<5x10^-8^ or P<0.05 thresholds. Y-axis is presented as proportion. Abbreviations: ‘IIa’ = FLL type IIa; ‘IIb’ = FLL type IIb; ‘III’ = FLL type III; ‘IV’ = FLL type IV.

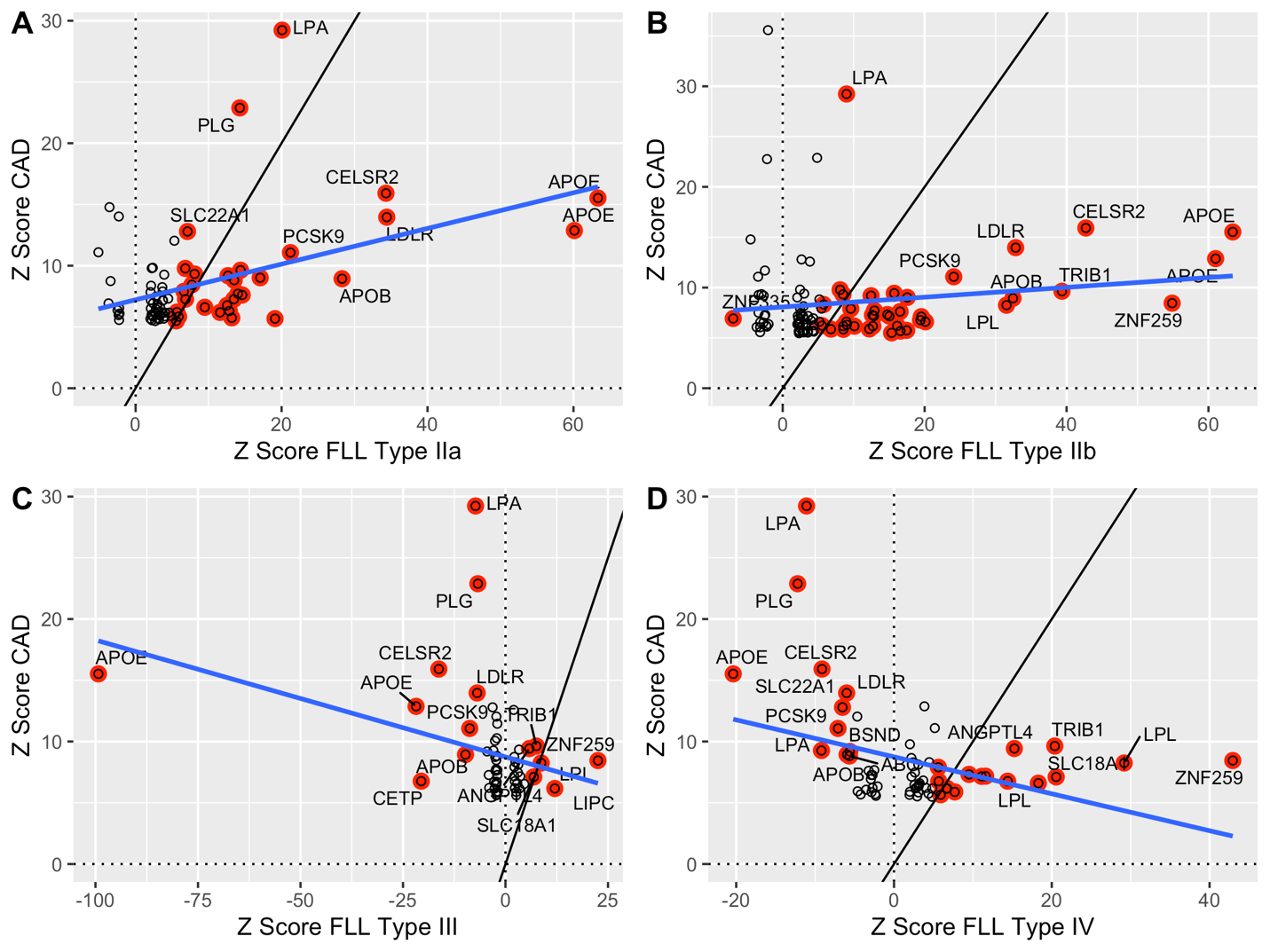

**Supplementary Figure 9. Scatterplots comparing Z-score for top CAD loci against the corresponding Z-score in each FLL GWAS.** For each phenotype (Type IIa – A; Type IIb – B; Type III – C; Type IV – D), 237 available SNPs were extracted from FLL GWAS summary statistics based on 241 independent CAD loci of the latest CARDIoGRAMplusC4D Consortium GWAS were extracted. For each FLL phenotype, all SNPs meeting nominal P-value <0.05 were retained for plotting. SNPs meeting genome-wide significance (P<5x10^-8^) in FLL GWAS are highlighted in red. Black line is x=y. Blue line is a linear model reflecting the relationship between x = Z Score FLL phenotype and y = Z Score CAD. Selected SNPs are annotated according to the nearest genes from the CAD GWAS.

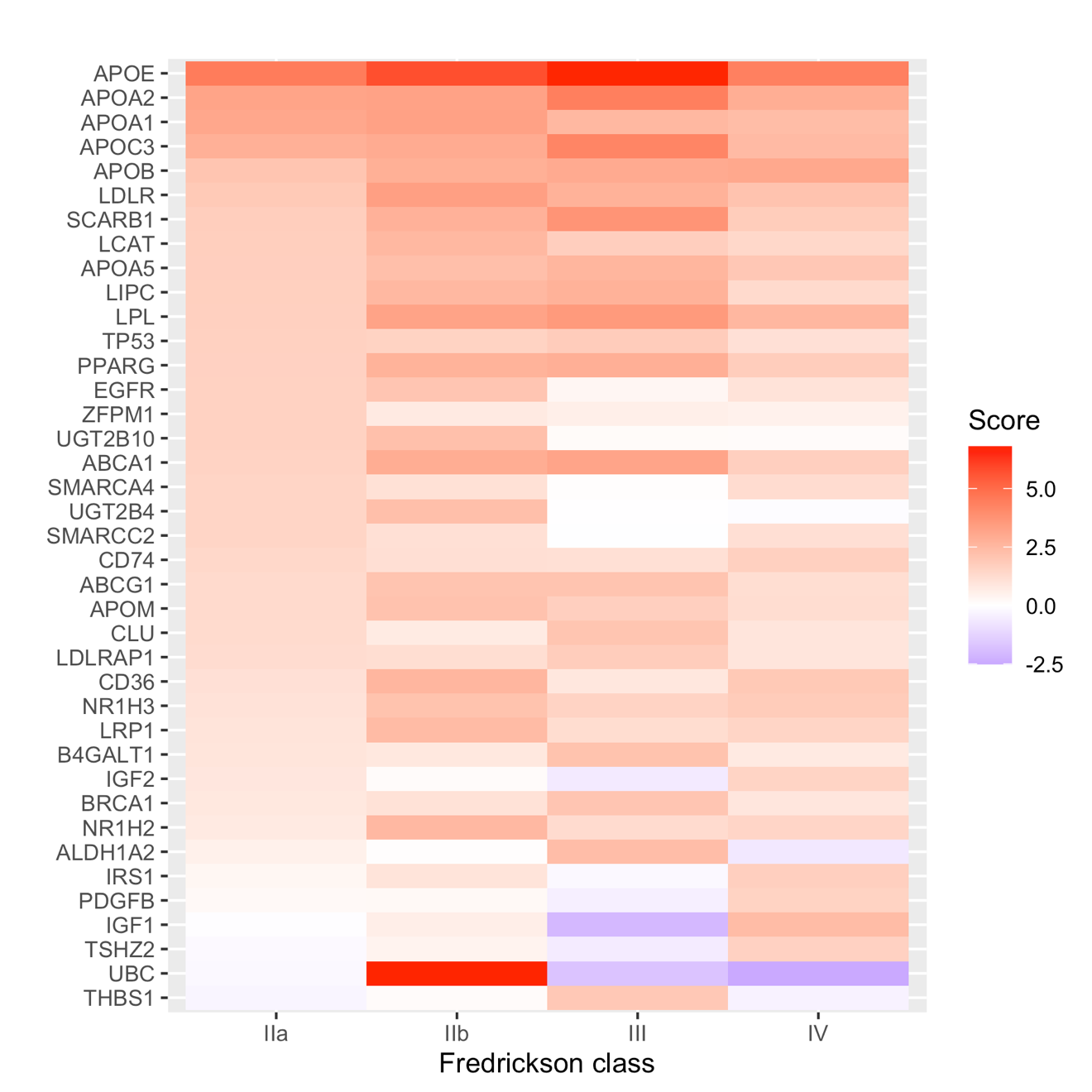

**Supplementary Figure 10. Polygenic priority scoring of top protein-coding loci contributing to each of FLL classes IIa, IIb, III, and IV.** The union of the top 20 PoPS scores was collated and is ordered according to PoPS rank for FLL type IIa. Cell coloring is according to absolute PoPS score, which reflects likelihood of contribution to each phenotype. Shades of red are likely contributors to a given phenotype, and shades of purple are unlikely to contribute significantly to a given phenotype.

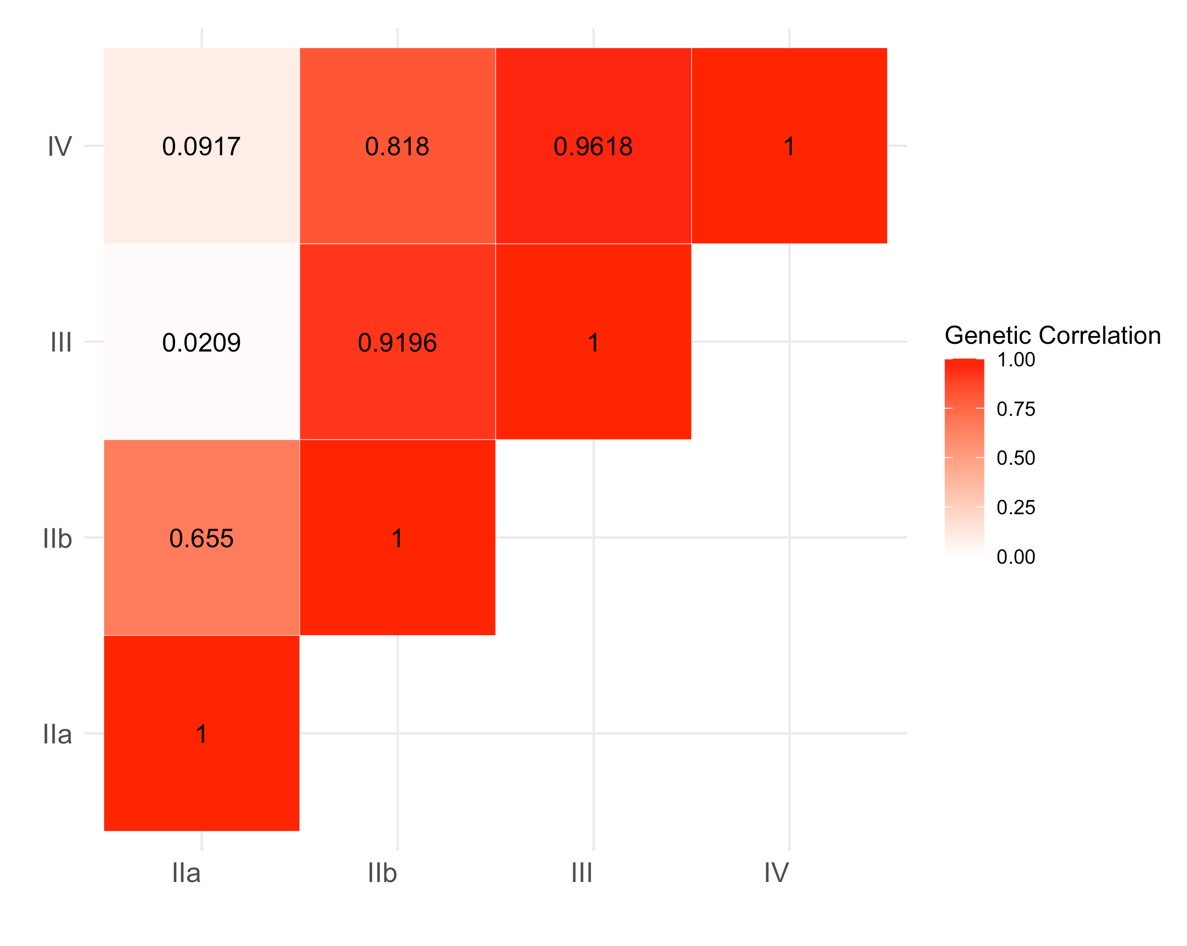

**Supplementary Figure 11. Genetic correlation between FLL phenotypes estimated with LD-score regression.** LD-score regression estimated the genetic correlation between FLL types IIa, IIb, III, and IV.

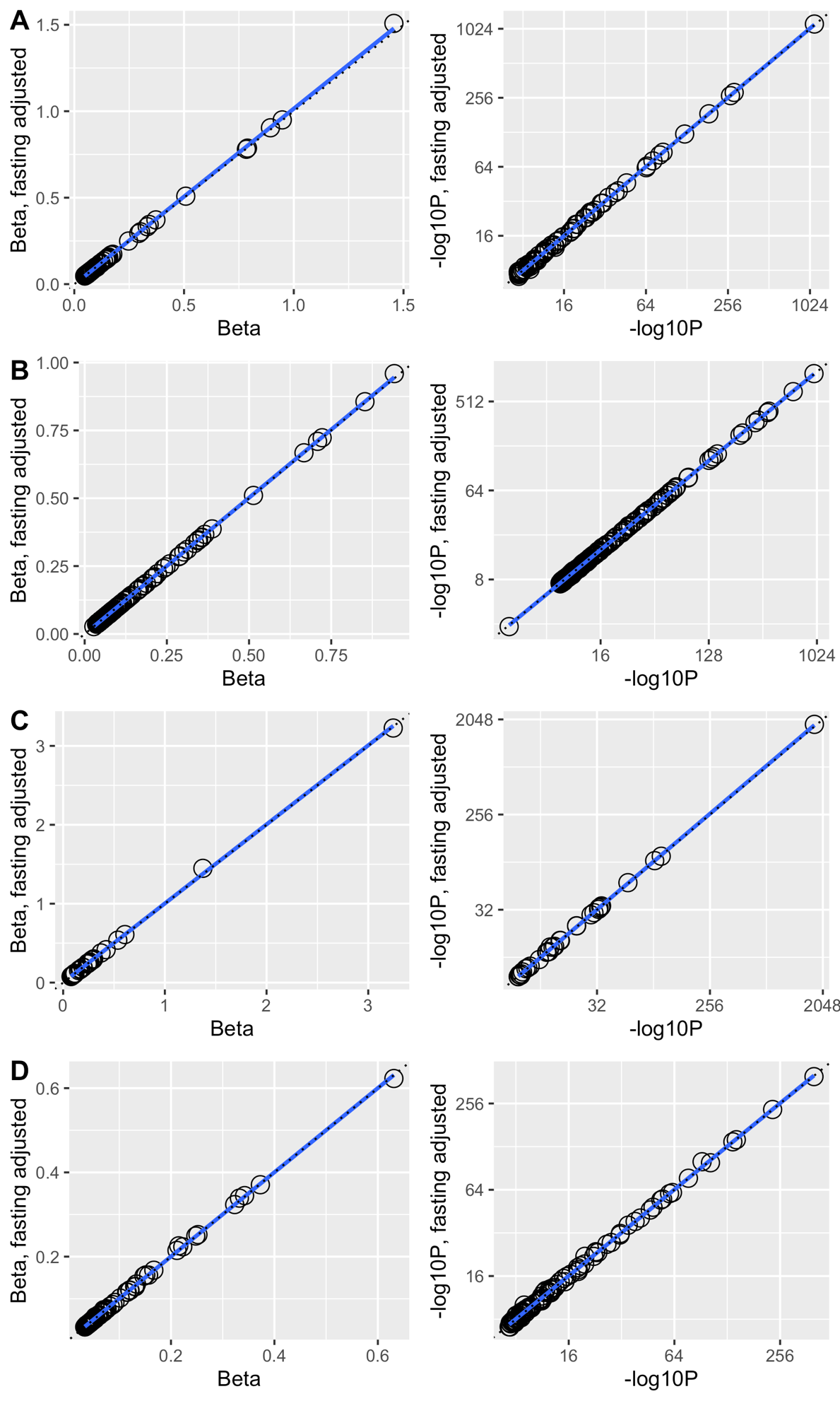

**Supplementary Figure 12. Comparing effect estimate and association P-value before and after adjustment for fasting time.** GWAS was repeated for top independent SNPs for each Fredrickson-Levy-Lees (FLL) phenotype after additionally adjusting for fasting time prior to blood draw. (A) FLL Type IIa; (B) FLL Type IIb; (C) FLL Type III; (D) FLL Type IV. For each phenotype, the left panel is the absolute value of the SNP effect estimate, and the right panel is the -log_10_ of the association P-value. The dotted line has slope = 1, Y-intercept = 0.
